# Ruling resistance out, and reading the blank: genome-based antimicrobial susceptibility prediction in clinical *Pseudomonas aeruginosa* isolates

**DOI:** 10.64898/2026.09.10.26362730

**Authors:** L. Scheper, J. Erdmann, A. K. Heroven, L. Knegendorf, K. Nielsen Leith, F. Boetius Hertz, S. Häussler

## Abstract

Genomic prediction of antimicrobial susceptibility aims to identify resistance determinants and thereby to report those antibiotics for which therapeutic failure is to be expected. We investigated what this approach can achieve in *Pseudomonas aeruginosa*, a species in which resistance often arises through changes in gene expression. For this purpose, 5,749 consecutive clinical isolates from the routine laboratories of two tertiary care centers were sequenced, each with paired phenotypic susceptibility test results for ciprofloxacin, ceftazidime, meropenem, and tobramycin.

Genomic variation could be interpreted only after three adaptations. Variants were called against the population-wide major allele rather than against PAO1. This removes markers that are artefacts of the reference. Loss of gene function was captured systematically, as many genetic determinants of resistance act through loss-of-function mechanisms. Furthermore, we introduced a specificity ratio — enabled by testing all four antibiotics on the same isolates — which separated markers associated with resistance mechanisms from those that merely tracked clonal lineage; without this distinction, expanding the catalogue reduced rather than improved predictive performance. In addition to classical resistance determinants, markers of susceptibility were taken into account, such as the functional loss of an efflux pump that exports the antibiotic out of the cell.

While resistance could only be assigned with corresponding confidence for a few percent of isolates, the absence of ciprofloxacin and meropenem resistance could be reported for substantially more isolates, in a conservative deduplicated analysis even for 43% and 87%, respectively. The remaining isolates were left blank rather than forced into one of the two categories. The blank is not an absence of information: resistance rates within it lay between those of the two reported groups, and its isolates clustered around the clinical breakpoint. It is the genomic counterpart of a population the phenotype already recognizes and reports as susceptible, increased exposure or as an area of technical uncertainty.

For meropenem, the residual risk of a susceptibility finding already meets the accepted threshold of 3%, while for ciprofloxacin it approaches this threshold. For tobramycin, validation in a larger cohort, possibly specifically enriched for resistant isolates, is required before our results can be considered reliable. For ceftazidime, by contrast, it appears that additional markers need to be identified, in particular those associated with increased AmpC expression. Taken together, the present data show that, for two of the four antibiotics investigated here, clinically relevant findings can already be generated today: the genome allows the identification of an isolate group with no indication of resistance, marks a smaller group as resistant, and assigns the remaining isolates to an intermediate range close to the clinical breakpoint.

## Introduction

The genomic prediction of antimicrobial susceptibility is already used clinically for *Mycobacterium tuberculosis* and *Staphylococcus aureus* [1,2]. At present, sequencing an isolate is not faster than the culture-based methods that are routinely used for the identification and susceptibility testing of pathogenic organisms. But, in contrast to culture-based susceptibility testing, a genome can reveal the mechanisms of antibiotic resistance, and it also provides high-resolution genetic information for surveillance. Looking to the future, it is likely that sequencing will be used more broadly in routine procedures in clinical microbiology, including direct application to patient specimens. If culture-independent sequencing from clinical material becomes routine, its clinical utility will depend substantially on the underlying resistance catalogue.

Genomic prediction works best when resistance is caused by acquired resistance genes or by well-defined mutations in antibiotic target structures. These are genetic variations that can be read directly from sequence data. In many pathogens, however, the dominant mechanisms of resistance are more complex, and *Pseudomonas aeruginosa* is a particularly illustrative example of this [3,4]. In this species, resistance is largely driven by gene inactivation and associated functional and regulatory changes. The bacterium can overproduce an intrinsic beta-lactamase, upregulate an RND efflux pump, or stop producing a functional porin. A genome sequence shows which genes are present, but it cannot directly show how much of each protein the cell produces or whether the product is functional.

Whether a genome can replace culture-based susceptibility testing has been assessed twice by a EUCAST subcommittee, in 2017 and again in 2025; the current report finds the evidence strongest where a clinical breakpoint coincides with the epidemiological cut-off value, and identifies complex, non-acquired resistance mechanisms as the main remaining obstacle [5,6].

The interpretation of the bacterial genome sequence for the prediction of antibiotic resistance is essentially based on two strategies. The first approach relies on curated resistance catalogues that interpret genetic variations at a defined set of chromosomal loci. In *P. aeruginosa* this line of work began with a survey that related the resistome of 390 clinical isolates to their susceptibility for three agents [7] and with an explicit measurement of how far resistome and phenotype correlate [8]; the largest validation study of a curated catalogue to date included 204 isolates [9]. The second approach uses machine-learning methods to identify predictive markers directly from the data. The best predictions for resistance in clinical *P. aeruginosa* isolates were achieved through combinations of genomic and transcriptomic information [10], the latter of which cannot be provided by routine diagnostics. Despite their differences, both approaches face the same problem, which was recently quantified. Bacterial populations are highly structured, such that a variant enriched in a resistant lineage can appear predictive even if it plays no causal role in the emergence of resistance. If the bacterial population structure is ignored, predictive performance is overestimated, and even increasing the training dataset cannot solve the problem [11].

To investigate the clinical utility of genomic resistance prediction under routine conditions in diagnostics, we assembled a collection of 5,749 *P. aeruginosa* isolates from the clinical microbiology laboratories of two tertiary-care centers. The isolates were collected and sequenced consecutively, without selection according to phenotype, sample type, or hospital ward. Antibiotic susceptibility test results were available for ciprofloxacin, ceftazidime, meropenem, and tobramycin. The collection is notable not only because of its size, but also because of two specific characteristics. First, all four antibiotics were tested on the same isolates. This allows a marker that causally mediates resistance to a specific antibiotic to be distinguished from a lineage marker that frequently increases resistance to several antibiotics non-specifically. Second, because the collection is consecutive and not curated, it can be assumed that both the resistance frequencies and the mixture of the underlying mechanisms reflect what a diagnostic laboratory encounters in routine practice.

We consistently used lineage-blocked validation to investigate for which antibiotics and for which isolates the genome provides enough information to support a reliable prediction of susceptibility or resistance. The genome can answer the question of whether an antibiotic is likely to be effective for far more isolates than the question of whether it is likely to fail. It follows that a catalogue of genetic determinants is most useful when it is used to rule out resistance. If neither resistance nor susceptibility can be attributed to a clinical isolate with high confidence, we leave the report blank. That blank should not be read as an absence of clinically usable information: with respect to both resistance rates and minimum inhibitory concentrations, these isolates fall between the two reported groups, and they form a category of their own.

## Methods

All details, including the rationale for the specificity ratio and the complete specification of the features and validation, are provided in the Supplementary Methods.

### Collection and susceptibility testing

Every *P. aeruginosa* isolate obtained from patient specimens in the clinical microbiology laboratories of two tertiary care centers during the study period was sequenced, irrespective of phenotype, specimen type or ward. Centre A contributed 2,972 isolates from 2021 and 2022, tested by broth microdilution, and Centre B 2,777 isolates from 2019 to 2024, tested by disc diffusion. The results were categorized retrospectively and uniformly, based on version 16.0 of the EUCAST breakpoint tables [12]. Because the susceptibility breakpoint for ciprofloxacin, ceftazidime and tobramycin in *P. aeruginosa* was set at 0.001 mg/L, the category S is not assigned for these agents. The categorization is therefore consistently “resistant” versus “non-resistant”, with resistance breakpoints of > 0.5, > 8, > 8 and > 2 mg/L for ciprofloxacin, ceftazidime, meropenem and tobramycin, respectively. Each result was additionally converted into a log2 distance from the breakpoint, such that MIC and inhibition-zone results share a common scale. Positive values indicate resistance.

DNA extraction from the clinical isolates, as well as Hackflex library preparation and 150-bp paired-end sequencing on NextSeq 500 or NovaSeq 6000 sequencing systems, were performed as described previously [13]. The reads were likewise trimmed, assembled and multilocus sequence typed using the same in-house workflow [13]. A total 5,749 genomes from isolates with a routine susceptibility result for at least one of the four antibiotics met the criteria for assembly and identification. Assembly quality was uniform (median 128 contigs, 99th percentile 350; minimum coverage 30-fold). The study was approved by the ethics committee of Hannover Medical School (No. 10372_BO_K_2022), the Regional Danish Patient Safety Authority (R-21015888), and the local data-protection authority (Pactius P-2020-743).

### Marker catalogue and features

Variants were called at 95 chromosomal loci. Seventy-six of them have a documented link to at least one of the four antibiotics and formed the candidate panels: 21 for ciprofloxacin, 47 for ceftazidime, 27 for meropenem and 18 for tobramycin. Nineteen loci have no documented link to any of the four agents; they were screened and yielded no marker that survived lineage control (Supplementary Note 6, Supplementary Table S1).

Four feature types were derived to assess de-functionalization of a protein: amino-acid substitutions, gene absence, frameshifts or premature stop codons in the variant annotation, and a shortening or lengthening of the respective locus by more than 5%. Positions are numbered according to the PAO1 reference sequence. Acquired resistance genes were identified with AMRFinderPlus [14] based on the gene name and substrate range; sequence types were assigned using the *P. aeruginosa* MLST scheme [15,16].

### Reference allele and marker blocks

Variants were determined relative to PAO1 and subsequently recoded relative to the allele carried by the majority of isolates at each position. In this way, the coding was reversed at 189 of 31,466 positions. Positions whose carrier sets overlapped with a Jaccard coefficient of at least 0.90 were merged into a single marker block.

### Determinants, specificity and validation

A marker block was considered a determinant if it was carried by at least ten isolates and at least half of these were resistant. Acquired enzymes were counted as determinants irrespective of the number of carriers and of the observed proportion of resistant carriers, since their contribution to resistance is established biochemically rather than empirically. The blocks were ranked according to the Wilson lower bound of their predictive value [17,18], and panels of 10 to 120 blocks were evaluated. Sensitivity and specificity are consistently reported for a fixed panel of 50 blocks. The antibiotic-specificity ratio is the odds ratio for one antibiotic divided by the geometric mean of the odds ratios for the other three antibiotics. A block is considered specific if this ratio exceeds 1.4 and the lower bound of its 95% confidence interval exceeds 1 (see Supplementary Methods).

In each cross-validation, complete sequence types were assigned to the folds, with marker ranking, block collapsing and the specificity filter recalculated within each training fold. The probabilities were derived from an L2-regularised logistic model based on seven features, and calibration was summarized using the Brier skill score against a baseline that takes only prevalence into account [19,20]. All analyses use the complete collection; the deduplicated sensitivity analysis (Supplementary Note 1) is reported wherever a clinical recommendation is made.

## Results

### A consecutive two-center collection

We analyzed a total of 5,749 *P. aeruginosa* isolates with high-quality draft genomes. For each of the four antibiotics, a susceptibility test result was available for at least 98% of the isolates, from between 2,828 and 2,898 patients. The collection is diverse: 5,081 isolates were distributed across 477 sequence types, with the largest accounting for 5.1%. The high-risk clones ST235 and ST111, at 1.1% each, ranked only 19th and 20th; the only high-risk clone among the twelve most frequent types is ST244 [21]. Resistance rates were 19.2% for ciprofloxacin, 13.9% for ceftazidime, 11.9% for meropenem and 8.3% for tobramycin, and were higher in Centre A than in Centre B for all four, most clearly for tobramycin (12.6% versus 3.7%). The two centers differ in several respects: Centre A contributed 2,972 isolates from 2021 and 2022, tested by broth microdilution, and Centre B 2,777 isolates from 2019 to 2024, tested by disc diffusion. Centre, susceptibility-testing method, sequencing batch, calendar period and patient population therefore cannot be compared directly (Supplementary Fig. 1).

### A population-aware reference removes pseudomarkers and reveals genuine gene loss

Using PAO1 as the reference, 66 of the 31,466 positions across the 95 loci were identified as variants in more than 90% of the isolates, and every isolate carried at least one such variant, including those isolates that were susceptible to antibiotics. One of these was *nalC* G71E, which was characterized in the PAO1 background and is listed as resistance-associated in curated catalogues [22]. However, it was present in 91% of the population. PAO1 accordingly carries the minority allele, and the apparent variant is the normal state of the species. Recoding each position against the majority allele removes these calls and proved more robust than filtering against a panel of susceptible reference strains, since this filter can exclude only those lineages that it originally contains (Supplementary Fig. 2, Supplementary Note 2).

A loss of the gene product was called as follows: complete absence of the gene, a disruptive frameshift or a premature stop codon, as well as shortening or lengthening of the gene relative to the population norm by more than 5%. Lengthening was rare in comparison, with 341 events in 339 isolates versus 9,463 shortenings in 5,667 isolates, and was concentrated almost entirely in *oprD* and *nfxB*. At both loci, the association with resistance was strong, but not antibiotic-specific and strongly confounded by lineage and acquired enzymes, such that neither was retained (Supplementary Note 2). The remaining loss-of-function calls were largely antibiotic-specific (Fig. 1) [23–28].

**Figure 1.**
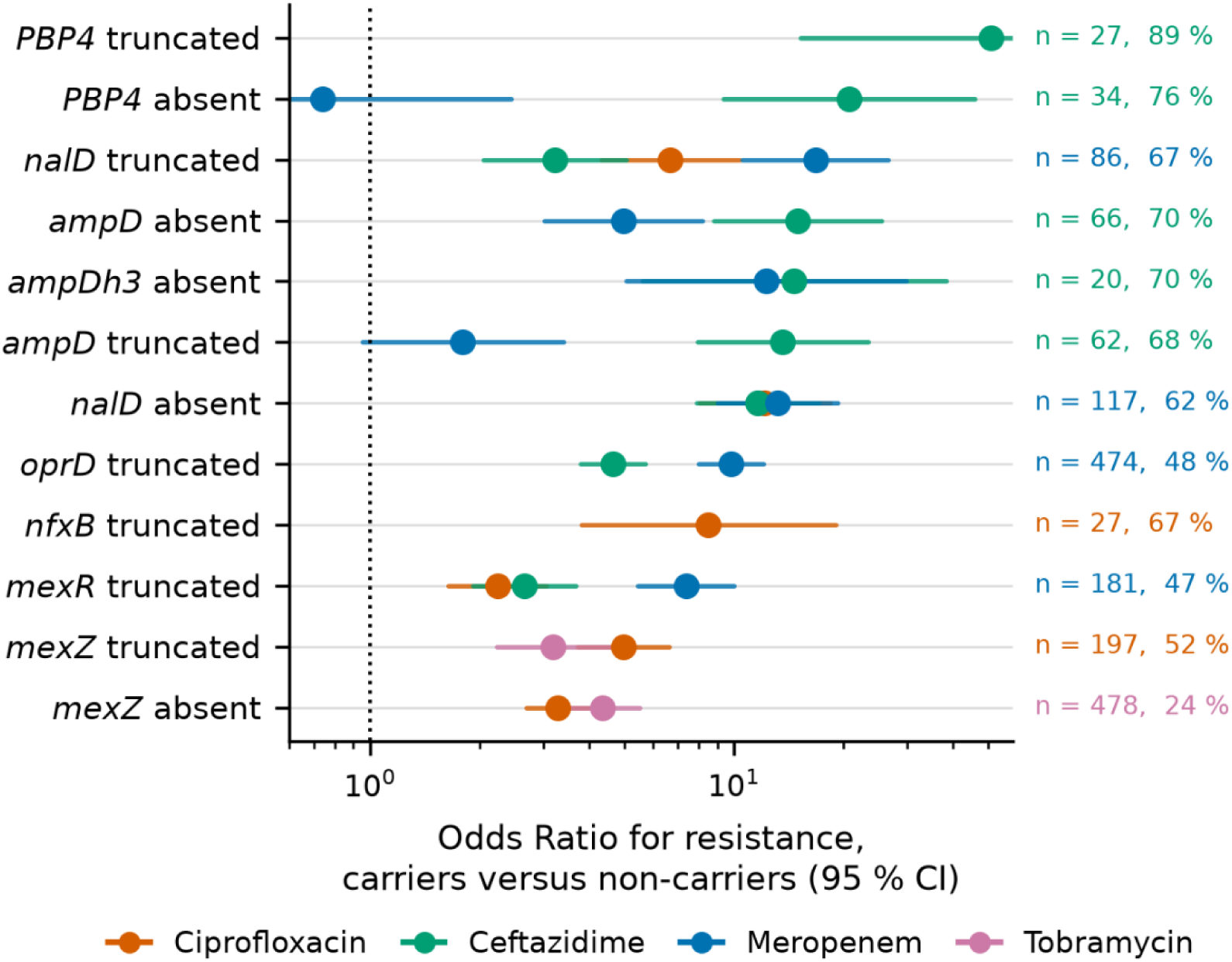
Loss of a gene product expands the catalogue by adding determinants. A gene product can be lost in three ways, and each was assessed: the gene can be absent, a frameshift or a premature stop codon can render the product non-functional, or the protein can differ in length. The odds ratios for resistance in carriers versus non-carriers of the respective determinants are shown, with 95% confidence intervals, for the different antibiotics in the same row, so that antibiotic specificity can be assessed directly. The number of carriers and the resistance rates are given for the antibiotic with the strongest association. The rows show the ten strongest associations overall, together with the two strongest associations for tobramycin, which is not represented among those ten.

Shortened PBP4 was the strongest chromosomal determinant in the dataset, with 89% of the 27 carriers resistant to ceftazidime, compared with 14% of non-carriers; the carriers were distributed across 14 patients and eleven sequence types. The signal therefore does not rest on a single clone, and after deduplication 16 carriers remained, of which 81% harboured resistant isolates.

### A criterion that separates mechanism from lineage

A variant that occurs frequently within a resistant lineage can appear highly predictive without having a mechanistic role [11]. Left in the catalogue, such a variant produces a resistance report for every isolate of that lineage, including the susceptible ones. Expanding the analysis from the 33 loci of previously characterized resistance determinants to all 76 introduced many blocks with high predictive value, but reduced cross-validated performance for three of the four antibiotics. We therefore defined the antibiotic-specificity ratio as the odds ratio for the antibiotic of interest divided by the geometric mean of the odds ratios for the other three. Across more than 1,800 marker blocks, 971 of 7,200 block–antibiotic pairs met the specificity criterion, and those that were retained are well-characterized resistance determinants: shortened PBP4, *gyrA*T83, *parC*S87, a premature stop codon in *mexR*, and *fusA1* substitutions at position 671 (Fig. 2) [29].

**Figure 2.**
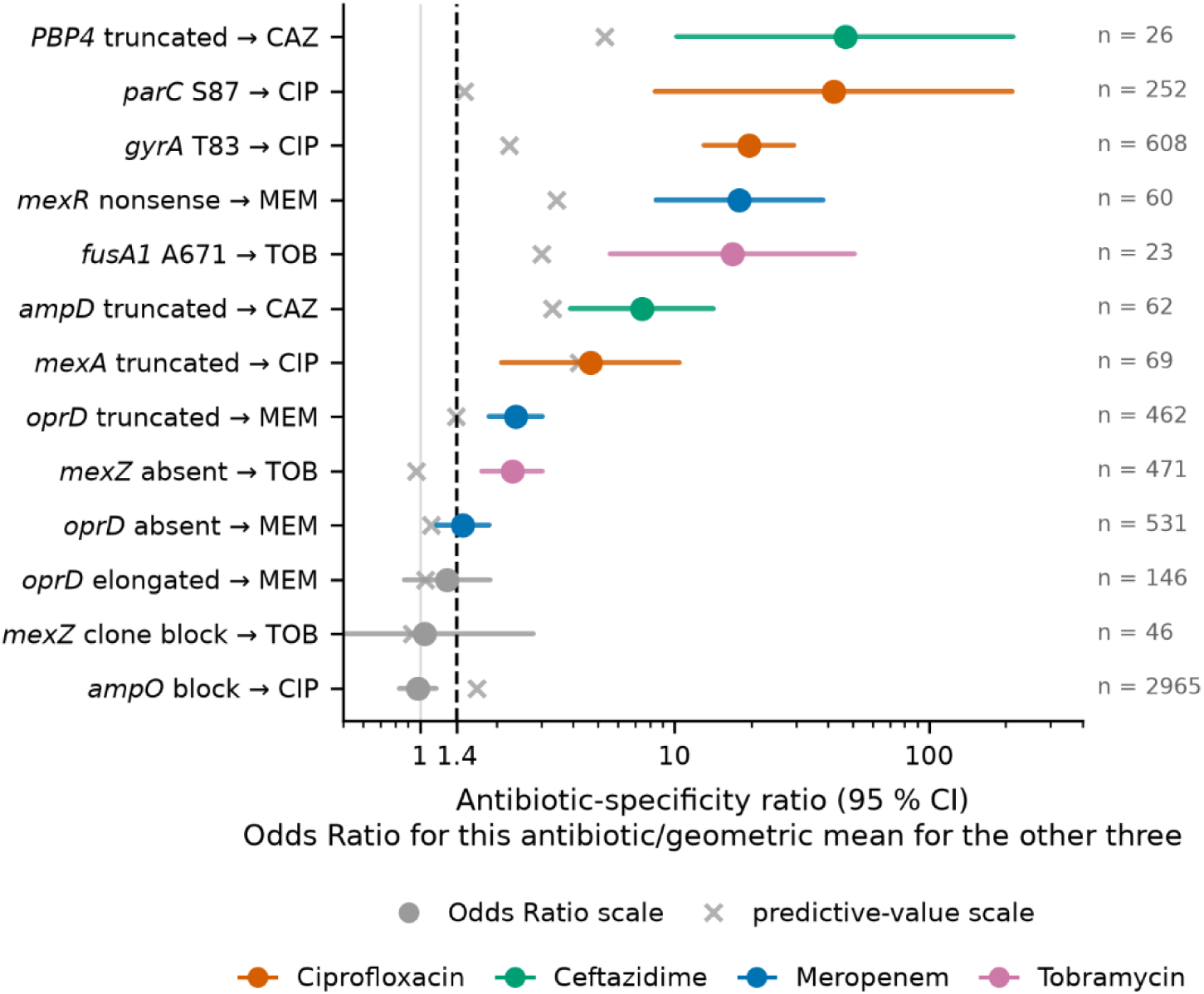
Causal determinants can be separated from lineage markers. A determinant that acts on one antibiotic is expected to predict resistance specifically to that antibiotic, whereas a marker of a multidrug-resistant lineage predicts resistance to several antibiotics non-specifically. The ratio that expresses this can only be formed because all four antibiotics were tested on the same isolates. Circles show the specificity ratios (the odds ratio for the antibiotic named in the row label, divided by the geometric mean of the odds ratios for the other three antibiotics), with their 95% confidence intervals; crosses show the same ratio formed on the predictive-value scale. A block is called antibiotic-specific when its odds ratio-based specificity ratio exceeds 1.4 and the lower bound of its confidence interval exceeds 1; in this case, it is shown in the colour of the respective antibiotic, and grey otherwise. Carrier numbers are those with a valid susceptibility result for the antibiotic named, so they differ slightly from Figure 1.

What was removed could not have been removed by a standard association test. A *mexZ* block carried by 46 isolates had 84.8% tobramycin-resistant carriers and an odds ratio of 62.7, but odds ratios between 49 and 81 for the three other antibiotics, resulting in a specificity ratio of 1.05; 39 of the 46 belonged to one sequence type, and 93.5% were resistant to ceftazidime, which is not exported by MexXY. The block therefore marks a lineage rather than the loss of *mexZ* function. The absence of *mexZ*, by contrast, was found in 471 isolates, reached a specificity ratio of 2.30 and was retained. Furthermore, considering predictive values alone would have discarded truncated *oprD*, the best-documented meropenem determinant in this organism, and retained an *ampO* block that reached a predictive-value ratio of 1.68, but only 0.98 on the odds-ratio scale. Its 2,965 carriers are no more likely to be ciprofloxacin-resistant than any other isolate.

Enlarging the catalogue without a correction of this kind is a step backwards. Removing the specificity filter reduced the Brier skill score for ciprofloxacin from 0.535 to 0.499 and for ceftazidime from 0.250 to 0.210. The deterioration in prediction is most visible in the resistance report: Without the filter, only 59.6% of isolates labelled meropenem-resistant were actually resistant, compared with 67.2% with the filter, and 72.8% compared with 81.4% for ceftazidime. More markers mean more variants that mark a lineage rather than a mechanism; a catalogue that grows without having a way to distinguish the two therefore becomes less rather than more reliable.

However, the criterion also penalizes mechanisms that are polyspecific: Of the 137 determinant blocks at the 33 mechanistically established loci, 98 (72%) met this criterion, with the highest retention for meropenem (93%) and the lowest for ceftazidime (54%). Where a mechanism acts on several of the antibiotics, mechanistic knowledge must take precedence over the empirical filter, and we applied this override twice: once for the loss of *nalD*, which derepresses MexAB-OprM and therefore raises the odds of resistance to three agents at once, and once for the structural genes of MexAB-OprM, which enter the model as a signed feature rather than through the filter (Supplementary Notes 3 and 4).

### Predictability follows the resistance mechanism

All four antibiotics were evaluated on the same isolates with the same variant detection, the same reference allele, the same validation and the same rule for panel construction, so that differences between them cannot be attributed to sequencing quality, population structure or the pipeline. Nevertheless, in a 50-block panel, cross-validated sensitivity and specificity ranged from 81.8% and 86.9% for ciprofloxacin down to 42.3% and 84.4% for tobramycin (Supplementary Table S2). The Brier skill score against a baseline that takes only prevalence into account was 0.535 (95% CI 0.467–0.602) for ciprofloxacin, 0.250 (0.163–0.354) for ceftazidime, 0.230 (0.134–0.328) for meropenem and 0.019 (−0.040 to 0.074) for tobramycin. For tobramycin, the interval includes zero. This means that the model essentially contains no information about tobramycin beyond the local resistance rate. The only exception is the rule-in pathway described below, since an acquired aminoglycoside-modifying enzyme identifies resistance, independently of what the model does.

The number of determinant blocks was 103 for ciprofloxacin, 156 for ceftazidime, 135 for meropenem and 46 for tobramycin. Ceftazidime therefore has the largest catalogue, but among the lowest sensitivity (Supplementary Fig. 3). A total of 155 ciprofloxacin-resistant isolates (14.1%), 230 ceftazidime-resistant isolates (29.0%), 143 meropenem-resistant isolates (21.4%) and 190 tobramycin-resistant isolates (40.1%) carried no detectable determinant at all.

A class of determinants is particularly useful when it is both precise, meaning that most of its carriers are resistant, and broad, meaning that it covers a large proportion of all resistance to that antibiotic (Fig. 3). Only ciprofloxacin has such a class: target-structure mutations in *gyrA* and *parC*, whose carriers were resistant in 84.1% of cases and which covered 74.7% of ciprofloxacin resistance. For ceftazidime, the acquired enzymes are precise but narrow, and the *ampC* pathway is broad but leaves four out of ten carriers susceptible. For meropenem and tobramycin, the broadest class is efflux regulation, which covers 49.8% and 44.7% of resistance, respectively, at predictive values of 61.1% and 64.2%.

**Figure 3.**
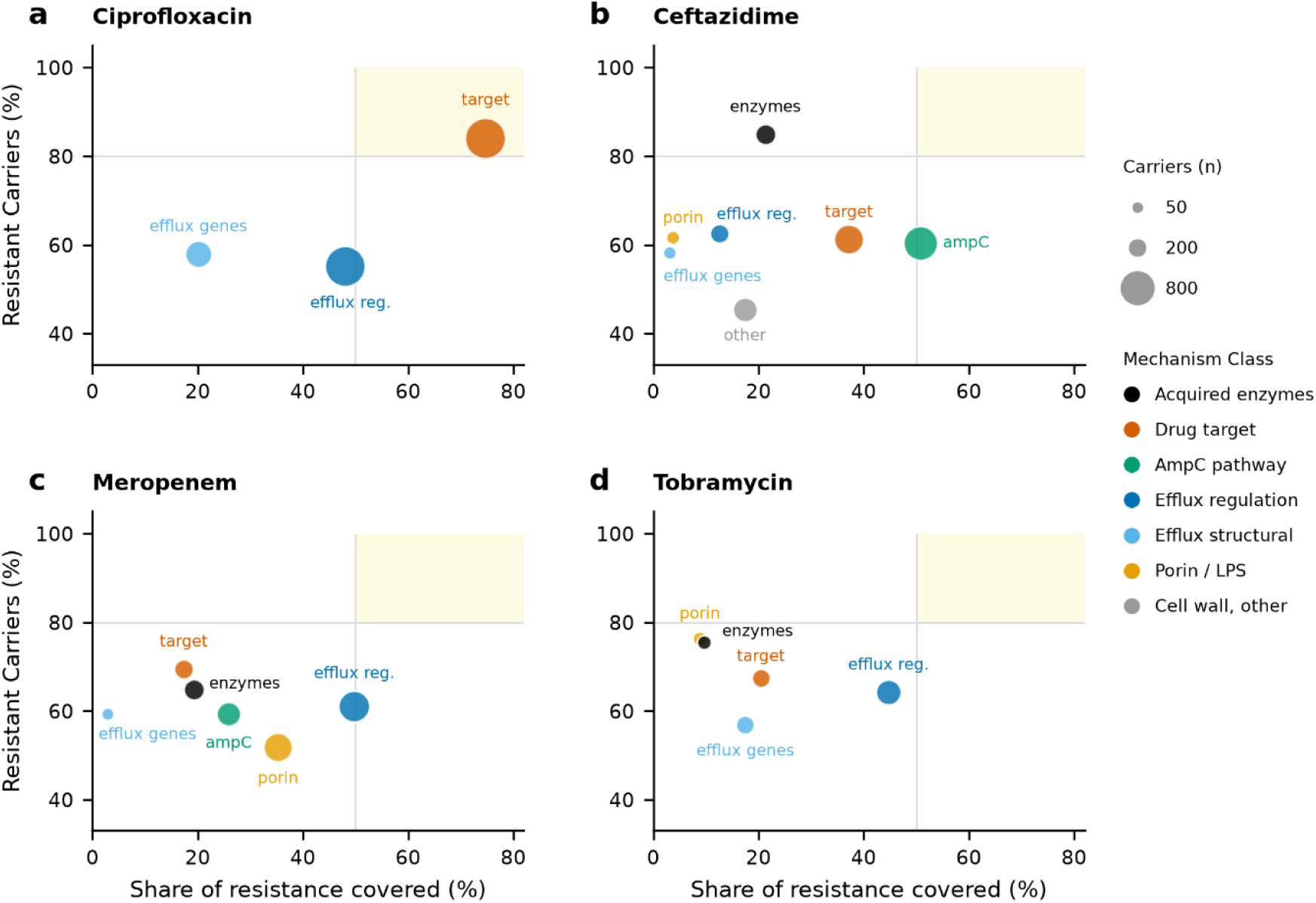
Predictability follows the resistance mechanism. The determinants of antibiotic resistance to (a) ciprofloxacin, (b) ceftazidime, (c) meropenem and (d) tobramycin were each assigned to one mechanism class according to the function of the locus they sit in, and each class was plotted with its positive predictive value (the proportion of its carriers that are actually resistant) against the share of resistance it covers (the proportion of all isolates resistant to the respective antibiotic that carry a determinant from this class). The area of the symbols is proportional to the number of carriers. Coverage does not sum to 100%, because classes overlap: an isolate can carry determinants from several classes. The shaded corner marks the range that a class would have to reach simultaneously in order to be both precise and broad.

### Loss of the exporting pump predicts susceptibility to meropenem and tobramycin

A pump that exports three of the four agents cannot produce an antibiotic-specific marker, so the structural genes of MexAB-OprM entered the model on mechanistic grounds rather than through the filter — the second of the two overrides. What they mark turns out to be not resistance but susceptibility. Because the structural genes of all four major RND efflux systems are included in the catalogue, the consequence of the loss of each pump for each antibiotic can be read out directly [30]. We examined all sixteen pump–antibiotic combinations; seven associations survived Bonferroni correction, and two of these showed increased susceptibility after inactivation of the structural genes of the efflux pumps. In both cases, the pump is known to export the corresponding antibiotic: MexAB-OprM for meropenem (stratified odds ratio 0.32) and MexXY for tobramycin (0.33) (Fig. 4a). The other five are discussed in Supplementary Note 4.

**Figure 4.**
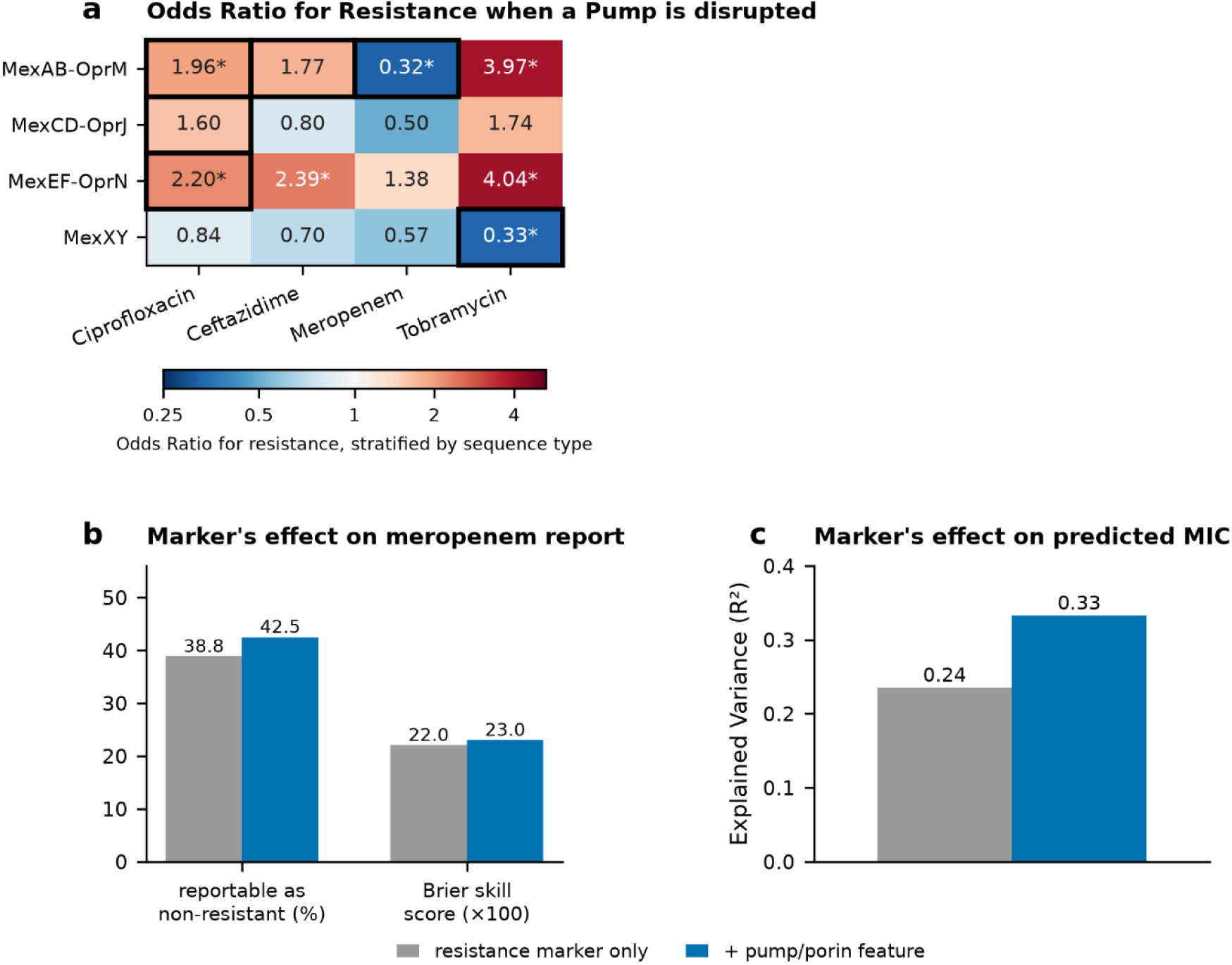
Loss of the exporting pump predicts susceptibility to meropenem and tobramycin. The structural genes of the four major RND efflux systems are part of the catalogue, so the effect of losing each pump can be read off directly for all four antibiotics. A pump is considered disrupted if the corresponding gene is absent, or if its protein product is truncated (due to a frameshift or premature stop codon) or atypical in length. Four efflux pump systems and four antibiotics give 16 pump–antibiotic combinations. (a) Mantel–Haenszel odds ratio for resistance in isolates with a disrupted versus an intact pump, stratified by sequence type so that the association cannot be driven by a single lineage; the strata themselves are not shown, only the pooled estimate. Blue (odds ratio below 1) indicates that carriers of the disruption are resistant less often than non-carriers; red (odds ratio above 1) indicates the opposite. Asterisks mark the seven combinations significant at the Bonferroni-corrected level of 0.05/16 = 0.003. Black outlines indicate antibiotics targeted by the respective efflux pump. The five combinations pointing towards higher resistance are discussed in Supplementary Note 4. The two protective combinations each involve the pump that exports the corresponding antibiotic: MexAB-OprM for meropenem and MexXY for tobramycin. (b) Adding a signed feature for pump disruption in isolates with an intact porin enlarges the group reportable as non-resistant without increasing its residual risk. The feature was tested for all four antibiotics and changed nothing for the other three. (c) The same feature applied to the MIC rather than to the category, in Centre A, where graded MIC values are available: explained variance of the log2 MIC rises from 0.24 to 0.33.

Of 71 isolates with shortened *mexA*, only one was meropenem-resistant, compared with 12% of the remaining isolates (odds ratio 0.11; 95% CI 0.01–0.76); stratification by sequence type shifted the odds ratio to 0.05, the carriers came from 38 patients and 28 sequence types, and after deduplication 40 remained, none of them resistant. Among the 1,019 isolates that carried an *oprD* defect, the pump remained protective, with meropenem-resistance rates of 43.2% when it was intact and 14.3% when it was not (p = 5 × 10⁻⁸). The protection decreased the more severe the porin defect was, falling to a relative risk of 0.57 when the gene was completely absent. This was expected, since in the absence of the porin, little drug enters the cell that could be pumped out again.

Including a signed feature for pump defect in isolates with an intact porin increased the Brier skill score of the model by a small but consistent amount (paired difference +0.010; 95% CI +0.005 to +0.016) and enlarged the group that could be reported as non-resistant. Applied to the MIC rather than to the category, it increased the explained variance of the log2 MIC from 0.24 to 0.33 (Fig. 4b, c). MexXY and tobramycin behaved similarly: among 362 isolates with functional loss of MexXY, distributed across 245 patients and 73 sequence types, 3.0% were resistant compared with 8.7% of the remainder (odds ratio 0.33), and 2.0% compared with 8.0% after excluding the 18 carriers of an acquired aminoglycoside-modifying enzyme.

### Marker detection is a graded function of the MIC

In Centre A, where MIC values were available, detection of a determinant increases steadily with the MIC (Fig. 5a): For the canonical *gyrA* and *parC* markers, it was 62% overall, but 91% among isolates with 4 mg/L or more (Fig. 5b). Isolates that carry a determinant and test susceptible have a much higher probability than non-carriers of lying within one twofold dilution step of the breakpoint (41% versus 11% for ciprofloxacin, 25% versus 4% for meropenem; p < 10⁻⁵ for all four antibiotics). The proportion of false-positive and false-negative isolates for the four antibiotics is shown in Fig. 5c.

**Figure 5.**
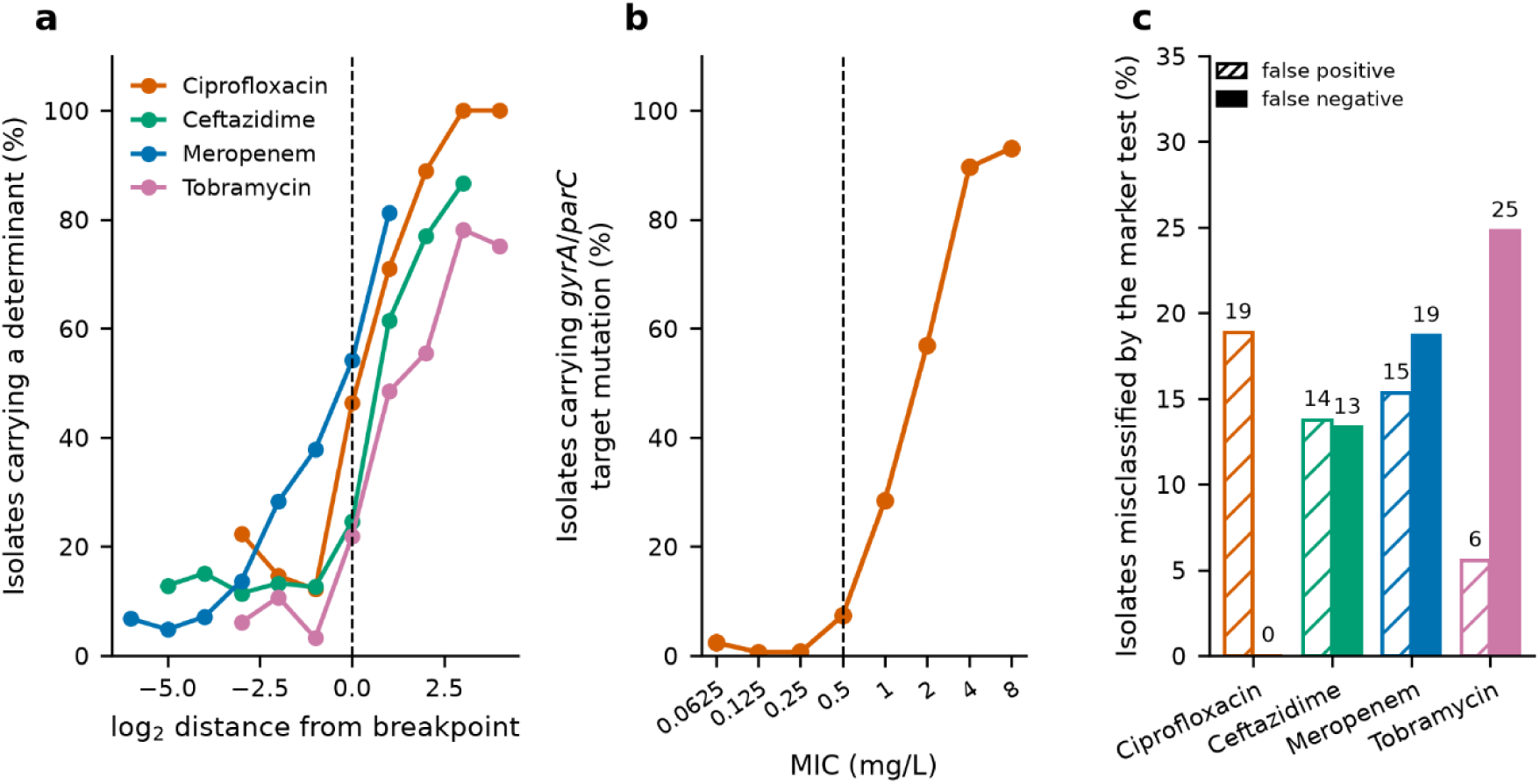
Marker detection is a graded function of the MIC. All three panels use data exclusively from Centre A, the only centre with graded MIC values. (a) Proportion of isolates carrying at least one determinant, plotted against the log2 distance from the clinical breakpoint for all four antibiotics. (b) The proportion of ciprofloxacin-resistant isolates in which a canonical gyrA or parC target mutation is detected is 62% overall but reaches 91% at an MIC of 4 mg/L or above. (c) Hatched: proportion of phenotypically susceptible isolates that nevertheless carry a determinant (false-positive result). Coloured: proportion of isolates at the highest measured MIC for the respective antibiotic in which no determinant is found (false-negative result).

For three of our four antibiotics, the EUCAST resistance breakpoint coincides with the epidemiological cut-off value [31,41]. The exception is meropenem. Its resistance breakpoint lies two doubling dilutions above the cut-off, at 8 against 2 mg/L. We repeated the analysis in Centre A with the endpoint set at the cut-off instead of at the clinical breakpoint. Predicting non-wild-type status raised the Brier skill score from 0.145 (95% CI 0.081–0.201) to 0.439 (0.356–0.530), and the sensitivity and specificity of a 50-block panel from 56.5% and 83.2% to 73.8% and 89.1%. Prevalence of resistance rose from 13.9% to 29.4%, and the group reportable as non-resistant shrank from 39.4% to 4.4% of isolates. The genome therefore describes the biological state of the isolate considerably better than it predicts the categorization as set by the clinical breakpoint — the same dependence on the alignment of breakpoint and cut-off that the current EUCAST assessment identifies as decisive [6]. A single categorical answer thus discards part of what the genome knows about the isolate, and we translated the model output into a three-state report instead.

### Excluding resistance reaches far more isolates than confirming resistance

For each isolate–antibiotic combination, the model outputs a predicted probability of resistance, and this allows three categories to be reported: isolates with a probability of being resistant of less than 5%, those with a probability of being resistant of more than 80%, and a blank in between. The lower threshold is anchored at the conventional limit of 3% for very-major errors [32]; the upper one follows no external convention, making it the more preliminary of the two (Supplementary Note 5).

Meropenem met the 3% requirement: among the isolates below the lower threshold, 1.9% were actually resistant (95% CI 1.4–2.5). Ciprofloxacin did not meet it in the complete dataset, with 3.6% (2.9–4.5), although the deduplicated value falls to 2.4%; tobramycin is above the limit at 4.2% (3.6–5.0). The non-resistant ceftazidime group contains a total of 51 isolates, with an observed resistance rate of 3.9% whose confidence interval ranges from 1.1% to 13.2% and is therefore too wide to act on (Table 1, Fig. 6b). Measured against the prevalence in the collection, resistance in this group runs 5.3-fold below it for ciprofloxacin and 6.3-fold for meropenem, but only 3.6-fold for ceftazidime and 2.0-fold for tobramycin.

**Table 1.** The three-state report, cross-validated.

|  | Ciprofloxacin | Meropenem | Ceftazidime | Tobramycin |
| --- | --- | --- | --- | --- |
| Resistance in the collection | 19.2 % | 11.9 % | 13.9 % | 8.3 % |
| Brier skill score | 0.535 | 0.230 | 0.250 | 0.019 |
| Reported non-resistant: $P(R) < 5\%$ | 34.3 % | 42.5 % | 0.9 % | 52.3 % |
| observed resistance there | 3.6 % | 1.9 % | 3.9 % | 4.2 % |
| fold below prevalence | 5.3 | 6.3 | 3.6 | 2.0 |
| Reported resistant, model probability only | 6.8 % | 1.0 % | 1.7 % | 0.2 % |
| observed resistance there | 88.5 % | 67.2 % | 81.4 % | 14.3 % |
| Reported resistant, model or enzyme | 6.8 % | 2.7 % | 3.2 % | 1.2 % |
| observed resistance there | 88.5 % | 87.6 % | 85.8 % | 60.3 % |
| Left blank | 58.9 % | 54.8 % | 95.9 % | 46.5 % |
| observed resistance there | 20.3 % | 15.9 % | 11.6 % | 11.6 % |

**Figure 6.**
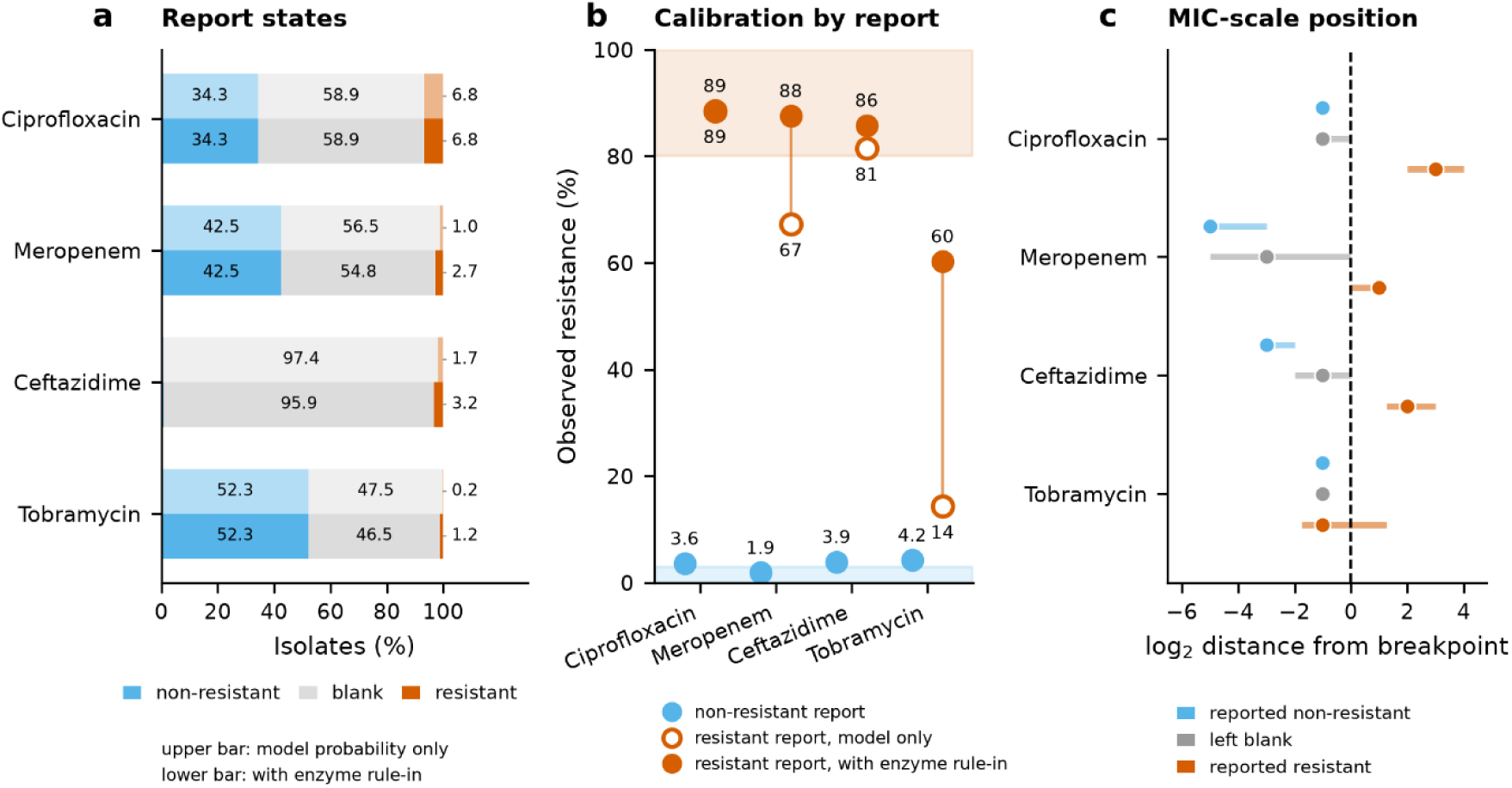
A report with three states. The report comprises three states—non-resistant, blank, resistant—reached via two independent routes into the resistant state: a calibrated model probability above 80%, or a documented acquired resistance enzyme. (a) Proportion of isolates in each state. The upper, pale bar uses the model probability alone; the lower, solid bar adds the enzyme rule-in, moving isolates from blank into resistant. (b) Observed resistance within each reported group against the two thresholds the report must meet. Blue circles show the resistant isolates assigned to non-resistant, which should lie within the shaded 3% very major error limit; only meropenem (1.9%) meets this. Open circles show the resistant group from the model probability alone; filled circles show it after the enzyme rule-in. The shaded area above 80% marks the precision that this group is intended to reach. Adding the enzyme-based route enlarges the group and moves it further into this shaded area, with no change for ciprofloxacin, for which the rule-in list is empty. The non-resistant ceftazidime group contains only 51 isolates, so its rate remains uncertain (95% CI 1.1–13.2%). (c) Median and interquartile range of the MIC (log2 distance from breakpoint, Centre A only) for the three reported groups, showing where each group sits relative to the breakpoint independent of how it was assigned.

An isolate carrying a metallo-beta-lactamase is resistant to meropenem for a biochemically established reason, independently of the frequency of this enzyme. An a priori-defined list based on the substrate specificities of the enzyme classes identified 111 isolates for meropenem, all of them resistant; 140 for ceftazidime, of which 94.3% were resistant; and 55 for tobramycin, of which 72.7% were resistant. These accounted for 16.6%, 16.6% and 8.4% of all resistance to the corresponding antibiotics, respectively. The model had not considered most of them (Supplementary Note 5).

Using both pathways together, model prediction and testing for the presence of acquired enzymes, made the resistance report for meropenem and ceftazidime both larger and more precise (Fig. 6a,b, Table 1). A definitive statement was then possible for 54% of the tobramycin predictions, 45% of the meropenem predictions, 41% of the ciprofloxacin predictions and 4% of the ceftazidime predictions, although the number for tobramycin reflects coverage rather than information. For ceftazidime, the small reportable proportion came almost entirely from rule-in. Throughout, the predicted probability depends on the local prevalence, and the model cannot be transferred between centers without recalibration.

Placed side by side, the two directions of the report are clearly unequal in their reach and very similar in their strength. The non-resistant report covered a third of the ciprofloxacin isolates, two fifths of the meropenem isolates and half of the tobramycin isolates, but under one per cent of the ceftazidime isolates; the resistant report consistently covered only a few percent (Table 1). Per isolate, however, neither direction is more informative: For meropenem, resistance in the non-resistant group was 6.3-fold below the prevalence and in the resistant group 7.4-fold above it, and for ciprofloxacin the factors were 5.3 and 4.6. The asymmetry therefore concerns coverage and not confidence.

### The third status of the result is not an absence of information

The observed resistance in the blank lay between the non-resistant and the resistant result for every antibiotic, and above the prevalence in the collection for three of the four (Table 1). The statuses non-resistant, blank and resistant are therefore ordered. In Centre A, where graded MIC values allow the position of a group on the dilution scale to be read directly, the same order holds for ciprofloxacin, ceftazidime and meropenem. It does not hold for tobramycin, where the resistant report contains only fourteen isolates. The blank is also the status most strongly concentrated around the breakpoint; it includes 43.3% of its meropenem isolates and 47.7% of its ceftazidime isolates within one twofold dilution step of it, compared with 8.1% and 14.3%, respectively, in the corresponding non-resistant results.

## Discussion

Our collection comprises 5,749 consecutively sequenced clinical *P. aeruginosa* isolates with paired susceptibility testing results for four antibiotics, thus more than twenty-five times the size of the largest knowledge-based validation in this species so far [9]. At that size, the focus can shift from average predictive performance to the specific isolates and antibiotics for which the model can support reliable conclusions. In general, a resistance catalogue is generated in order to identify resistance. In this collection, however, it fulfilled the opposite task better: a genomic statement of non-resistance was possible for one third to two fifths of the ciprofloxacin and meropenem isolates and, in the conservative deduplicated analysis, for 43% and 87% of these isolates, respectively. A statement of resistance, by contrast, was possible only for a few percent.

Per isolate, the two statements are of comparable informative value. The asymmetry therefore lies not in predictive performance, but in how often the respective statement can be made at all.

In principle, a categorical result requires that every isolate be assigned to one of two categories (resistant, non-resistant), even when the prediction is uncertain. But then there is no possibility for the clinician to recognize how robust this assignment is. We therefore introduced a blank whenever the genome cannot make a statement about resistant or non-resistant. But it is more than a blank. With regard to the observed resistance, the isolates contained in it lie between the two reported groups, and they cluster closer around the breakpoint than these groups. A blank is therefore itself a statement about the isolate. Such a population has previously already been described both by the EUCAST category “susceptible, increased exposure” and by the area of technical uncertainty [33,42].

For the meaningful interpretation of genomic variation, we introduced three corrections: the recoding of variants according to the population majority allele, whereby markers are removed that are artifacts of the reference; the assessment of the loss of a gene product, whereby determinants are captured that cannot be detected by substitution-based variant detection; as well as the specificity ratio, which distinguishes a marker of mechanism from a marker of lineage. The third criterion validates the markers again independently of deduplication by patient and sequence type. In this way, markers that are based on a few multidrug-resistant clones lose the greater part of their predictive value. The first two corrections follow from using a single reference strain and from scoring function rather than sequence, and are not specific to this species. The third requires collection in which several antibiotics have been tested on the same isolates — a constraint of study design, not of the data themselves.

The significance of the presence of an enzyme whose resistance-mediating activity has been demonstrated biochemically cannot be learned comprehensively from statistical models. We therefore introduced a rule-in rule on the basis of an established mechanism, as is usual in existing knowledge-based schemes [9]. Moreover, in addition to resistance determinants, we also considered markers of susceptibility. One such marker is the functional inactivation of an efflux pump, which is not uncommon.

Our predictive performance is lower than previously reported. For ceftazidime, our sensitivity was 43.4%, compared with 72.4% (54-85) reported by Cortés-Lara and colleagues [9]. Their estimate was based on 29 resistant isolates, and discordant isolates were reassessed by repeated MIC testing and RT-PCR, with adjustments made where appropriate. Their figure therefore reflects agreement with a curated reference phenotype, whereas ours reflects results generated by routine diagnostic laboratories. Their filter for natural polymorphisms also excluded any variant identified in a susceptible strain. This improves specificity, but by definition it removes determinants with incomplete penetrance. As a result, the intermediate population we describe could not have been identified within that framework.

Before WGS-based molecular diagnostics can be introduced into clinical practice, several obstacles still need to be addressed. Some of our strongest signals are still based on only a few lineages. Our filters reduce the confounding between mechanism and clone, but they do not eliminate it. The probability that a prediction is correct also depends on local prevalence. Accordingly, each laboratory requires a calibration dataset large enough to estimate the residual risk. In addition, predictive performance is currently defined in terms of categorical agreement [32]. This is not well aligned with a probabilistic output, even though the EUCAST area of technical uncertainty moves in a similar direction [33]. For the same reason, the endpoint itself is also open to question. A clinical breakpoint is not a property of the isolate. It marks the concentration up to which an antibiotic is expected to remain effective under an agreed dosing regimen. Non-wild-type status, by contrast, is a property of the isolate. Ideally, a catalogue should therefore be developed against the epidemiological cut-off, even if the laboratory report must follow the currently applicable clinical breakpoint.

Our results are intended to show what a genomic susceptibility report can achieve. For ciprofloxacin and meropenem, it identifies susceptibility in one third to two fifths of isolates, with a residual risk that reaches the accepted threshold for meropenem and approaches it for ciprofloxacin. For isolates for which no result is issued, it indicates that they fall between wild-type and clearly resistant isolates. For tobramycin, the reportable group is the largest of all, but the resistance rate within it lies only 2.0-fold below the local rate. Tobramycin has the smallest catalogue, with 46 determinant blocks, and two in five resistant isolates carry no detectable determinant. This points to a sample size problem that should be examined in larger collections specifically enriched for resistant isolates. Ceftazidime, by contrast, has the largest catalogue but the poorest predictive performance. What is missing here is a search for events that may increase intrinsic AmpC activity. Raised expression can arise from many different upstream changes, several of which may lie outside our loci or be individually too small to pass the detection threshold. Resistance mediated by altered gene expression may therefore stay systematically hard to catalogue.

For the clinical application of WGS-based molecular diagnostics, this does not mean that the road ahead is still very long, but rather that it leads in a different direction from what has previously been assumed: not toward a catalogue that identifies which antibiotic will fail, but toward a report that indicates susceptibility and explicitly marks uncertainty as such. In this direction, prediction for two of the four antibiotics tested here is already robust today. Both conclusions of the most recent EUCAST subcommittee assessment [6] are addressed by our results: predictive performance is strongest where the clinical breakpoint coincides with the epidemiological cut-off, and the main obstacle it identifies — complex, non-acquired resistance mechanisms — can be partly overcome by reporting explicitly where the genomic evidence is insufficient.

## Data Availability

All data produced are available online at github.com

https://github.com/LindaScheper/pseudomonas-gAST

## Declarations

### Ethics

The study was approved by the institutional review board of Hannover Medical School (No. 10372_BO_K_2022), which waived the need for informed consent. On the Danish side no ethical approval was required, the study being retrospective and non-interventional; permission to use routine laboratory records retrospectively was granted by the Regional Danish Patient Safety Authority (R-21015888) and the local data-protection authority (Pactius P-2020-743). The analysis uses no patient data. Each isolate enters it as a genome sequence, a routine susceptibility result and a pseudonymous identifier that links repeated isolates from the same person; no clinical, demographic or identifying information are used in this study.

### Data availability

Every genetic variant on which this study rests is released. The deposited files comprise the complete variant table for all 95 loci (listing every amino-acid substitution, truncation, frameshift, premature stop codon and gene-absence call per isolate) the acquired-resistance-gene calls, the derived marker matrices, the isolate table with centre, sequence type, pseudonymous patient identifier and susceptibility result for each antibiotic, the exact cross-validation fold assignment and the locus list. They are available at [https://github.com/LindaScheper/pseudomonas-gAST], together with the analysis and figure scripts, which reproduce every number and every figure in this article from those files.

## Author contributions

Data collection: J.E., L.K., K.L.N. F.B.H.; Project Design: S.H., L.S., J.E., AK. H.; Data Analysis and visualizations: L.S., J.E.; Manuscript writing: S.H., AK.H., L.S.; Proofreading and finalization of manuscript: S.H., AK.H., L.S.

The final version was approved by all authors.

## Use of AI assistance

The authors used Anthropic’s Claude as an analytical and editorial assistant: to write and check analysis code, to recompute reported quantities from the primary data, to draft and revise text, and to audit the manuscript for internal consistency between text, tables and figures. Every analysis was specified and inspected by the authors, who verified the reported results and take full responsibility for the content of this work.

## Figures

**Supplementary Figure 1.**
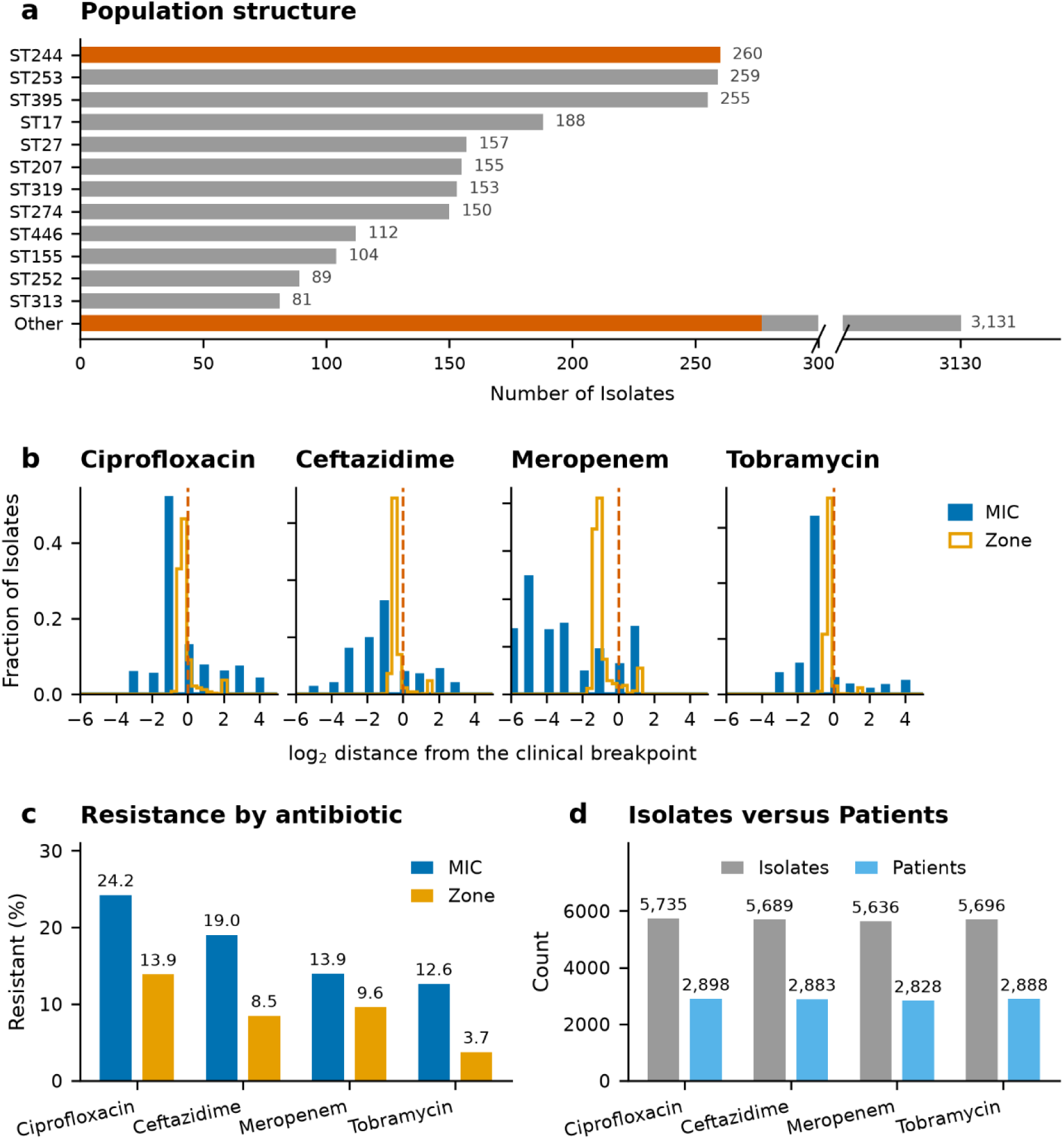
A consecutive two-centre collection. Every *P. aeruginosa* isolate recovered from a patient specimen during the study period was sequenced without selection; 5,749 had both a quality-controlled genome and an antibiotic susceptibility result. (a) Population structure shown for the 5,094 isolates with an assignable sequence type. The twelve most frequent sequence types; ST244, the only internationally recognised high-risk clone among them, is highlighted. (b) Distribution of resistance for the four antibiotics studied, expressed as the log2 distance from the current clinical EUCAST breakpoint, so that MIC and inhibition-zone results share a common scale. Zero marks the breakpoint; positive values indicate resistance. (c) Resistance rates by antibiotic and centre. Centre A has higher values for every antibiotic (d) Isolates in relation to patients: the collection contains serial isolates, and 30.6% of the isolates come from patients who contributed five or more isolates. A deduplicated sensitivity analysis is shown in Supplementary Note 1.

**Supplementary Figure 2.**
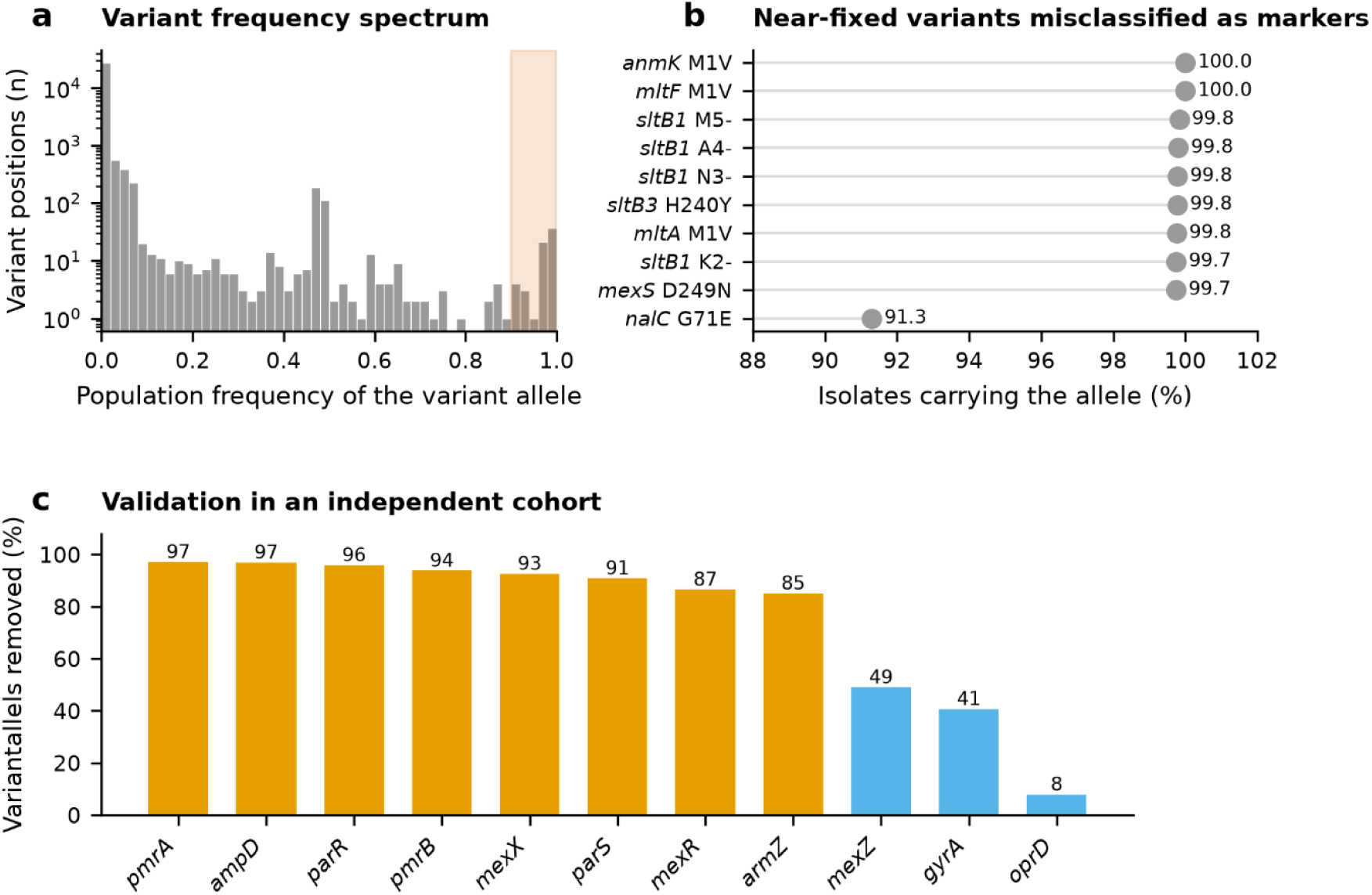
Reference-based variant calling systematically generates false markers. Calling variants against PAO1 generates pseudomarkers carried by nearly every isolate. (a) Frequency distribution of the variant allele across the called positions in the 95 curated loci. The shaded area marks positions at which more than 90% of the isolates carry the “variant”. At these positions, PAO1 carries the minority allele of the species. Sixty-six positions exceed a population frequency of 90%. (b) The ten most frequent pseudomarkers together with nalC G71E—a substitution described in the primary literature as resistance-associated and listed in curated catalogues on this basis, but present in 91% of this population. (c) Proportion of variant calls removed after filtering out natural polymorphisms, shown by locus, for loci that are also included in the collection of Cortés-Lara et al. In eight of the eleven loci, the proportion of removed variants exceeds 85%. These include the regulators *pmrA, pmrB, parR, parS, mexR* and *armZ*. The proportion is lower for *mexZ* (49%) and *gyrA* (41%), and negligible for *oprD* (8%), where the calls are genuine disruptions.

**Supplementary Figure 3.**
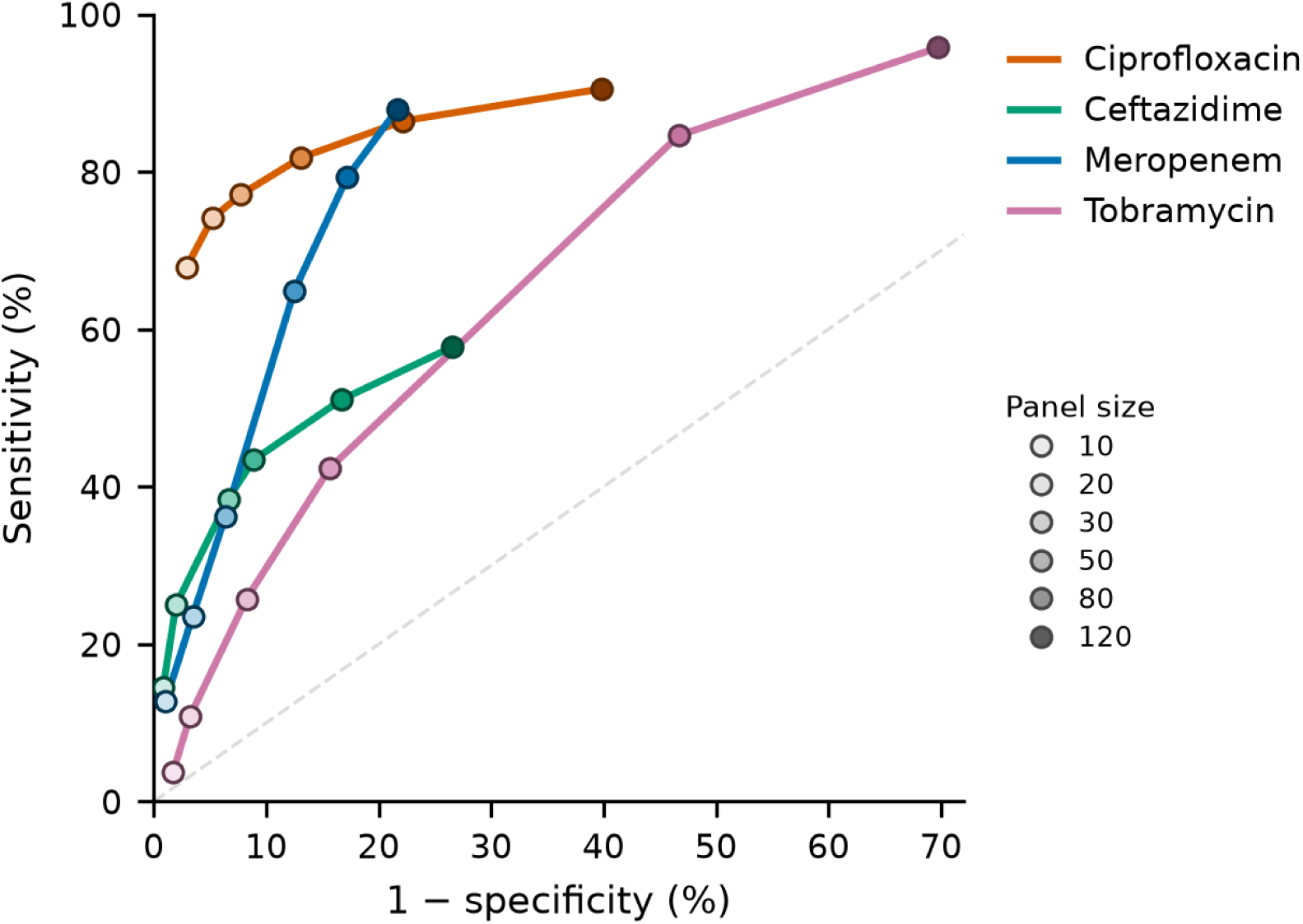
The ranking of the four antibiotics does not depend on panel size. Sensitivity is plotted against 1 − specificity for panels containing 10 to 120 marker blocks. Cross-validation was performed by assigning complete sequence types to individual folds. Point color indicates panel size, with darker shades denoting larger panels. The dashed diagonal represents chance performance, for which sensitivity equals 1 − specificity.

## Supplementary material

### Supplementary Methods

#### Sequencing, assembly and quality control

A genome was included in the analysis only if the sequencing and assembly quality controls had been passed and the isolate had been identified as *P. aeruginosa*. 63 genomes were excluded because 20 or more of the 95 catalogue loci were missing from the annotation; each would have contributed a spurious loss of the gene product at every one of them. Among the genomes retained, no isolate showed an unusually high number of missing catalogue loci.

#### The antibiotic-specificity ratio

To determine the antibiotic specificity of a marker, we compared its odds ratio for the antibiotic under investigation with the geometric mean of the odds ratios for the other three antibiotics. The contrast is calculated on the logarithmic scale, where the geometric mean corresponds to the arithmetic mean, and is then back-transformed; an arithmetic mean of the odds ratios themselves would produce a different, scale-dependent result. Because the null value is 1.00, a ratio was considered statistically different from the null when its 95% confidence interval excluded 1. The threshold of 1.4 serves only to exclude blocks with apparently insufficient specificity.

The threshold of 1.4 derives from a null experiment on the predictive-value scale. In this experiment, the positive predictive value for one antibiotic is set in relation to the geometric mean of the values for the other three antibiotics; a predictive value depends both on prevalence and on the odds ratio. Ciprofloxacin resistance is the most common in this collection (19.3% compared with 14.0% for ceftazidime, 11.8% for meropenem and 8.4% for tobramycin). Therefore, a marker with an identical odds ratio of 2 for all four antibiotics gives a ratio of 1.59 for ciprofloxacin, even though it has no preference whatsoever for a particular antibiotic. Depending on the assumed odds ratio, the null value lies between 1.11 and 1.59; 1.4 was set within this range. However, little depends on it: increasing the threshold from 1.0 to 1.4 removes 47 of 1,018 qualifying block–antibiotic pairs. No pair has a ratio between 1.0 and 1.2 with a lower confidence bound above 1, because a ratio so close to the null value cannot have a confidence interval that excludes it.

A Haldane correction was applied to the odds ratios. The confidence interval of the contrast was calculated by error propagation, treating the four outcomes as independent. This is conservative because they are positively correlated within isolates.

#### Validation and model

Cross-validation used five folds repeated three times, with complete sequence types assigned to the folds. Each isolate was therefore evaluated by a model trained without that isolate and without any other member of its lineage.

The model used seven features: three counts, namely the number of carried blocks among the top 12, 50 and 120 blocks in the ranking for the respective fold; three indicators for the presence of any acquired resistance gene, any truncation call and any gene-absence call; and one signed feature for disruption of the structural genes of MexAB-OprM in isolates with intact oprD, which was offered to all four antibiotics. These seven features constitute the primary model, and every performance figure reported in the main text and in Table 1 comes from it. The model carries a single signed-feature slot; replacing MexAB-OprM in that slot by a corresponding feature for MexXY disruption was evaluated separately for tobramycin and is reported only in Supplementary Note 5. MexXY is not part of the primary model.

The confidence intervals were determined by bootstrap resampling of complete sequence types.

#### Matched analysis

Associations were additionally estimated within sequence types, using the Mantel-Haenszel estimator over those sequence types that contain both resistant and susceptible isolates, with one isolate retained per patient and sequence type, and a Bonferroni threshold over the blocks tested (Supplementary Note 6). This holds the genetic background constant by construction rather than adjusting for it.

#### Serial isolates

30.6% of the isolates came from patients who contributed five or more isolates.

### Supplementary Note 1. Serial isolates inflate the estimates

Because patients with resistant infections may be more likely to undergo follow-up sampling, our collection may include several isolates from the same patient. We therefore repeated the entire analysis using one isolate per patient and sequence type, according to two selection rules: the chronologically first isolate from each group, following the CLSI M39 convention [34,43], and the most resistant isolate. Isolates without a patient identifier were retained individually. This deduplication reduced the collection from 5,735 to 3,045 ciprofloxacin-tested isolates, from 5,689 to 3,023 ceftazidime-tested isolates, from 5,636 to 2,974 meropenem-tested isolates, and from 5,696 to 3,029 tobramycin-tested isolates.

**Supplementary Note Figure 1.**
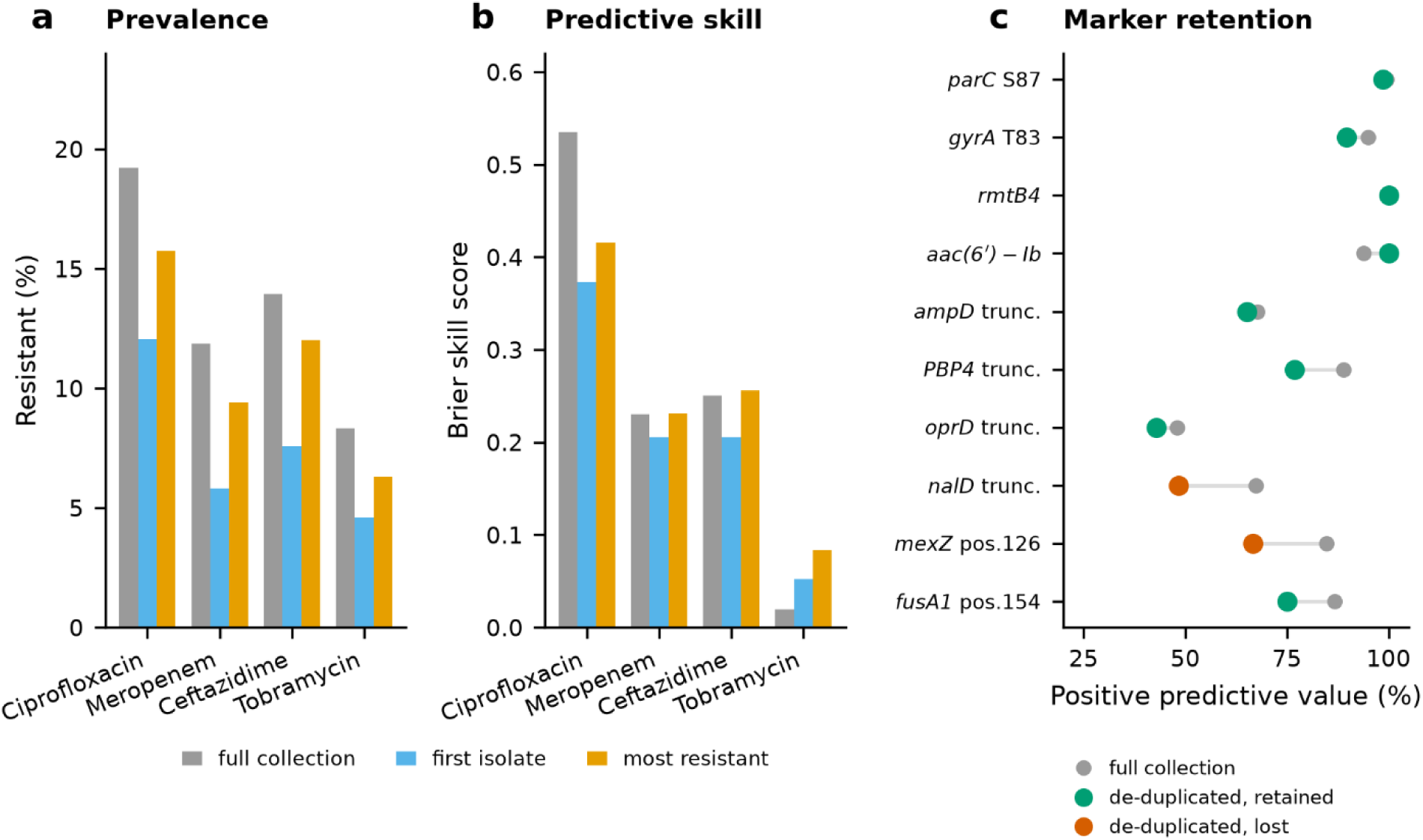
Serial isolates lead to an overestimation. A deduplicated sensitivity analysis with one isolate per patient and sequence type. Two selection rules were applied: the chronologically first isolate of each patient–sequence-type group, following the CLSI M39 convention, and the most resistant isolate of the group. **(a)** Resistance prevalence under the respective rule. **(b)** Brier skill score under the respective rule. Ciprofloxacin loses the most (0.54 → 0.37), ceftazidime and meropenem lose less (0.25 → 0.21 and 0.23 → 0.21), and tobramycin improves (0.02 → 0.05). **(c)** Positive predictive value of individual determinants before and after deduplication, under the first-isolate rule. Target-site mutations and acquired enzymes retain their value; the markers that lose predictive value are consistent with lineage-associated signals driven by repeated isolates from a few clones. Position 154 in *fusA1* falls from 87% to 50%—the same conclusion reached by the antibiotic-specificity ratio through an independent route.

The reason why ciprofloxacin loses the most is that many non-resistant isolates in the complete collection are clonal repeats from a small number of patients. As a result, a marker that these clones happen not to carry appears more specific than it actually is; after deduplication, it encounters a genuinely diverse population of susceptible isolates. Resistance prevalence also falls, from 19.2% to 12.1%, so that the group reportable as non-resistant grows from 34.3% to 43.3% of the isolates and the observed resistance within this group falls from 3.6% to 2.4%.

After deduplication, fewer blocks qualify as determinants, since in an approximately halved collection many fall below the minimum of ten carriers. Among the remaining blocks, the retention of predictive value depends on the type of marker. Target-site mutations and acquired enzymes remain almost unchanged (Supplementary Note Figure 1c).

We report the complete collection as the primary analysis because it reflects the diagnostic situation, in which a laboratory sequences the isolate in front of it and not an epidemiologically curated selection. Sequence-type-blocked cross-validation already assigns serial isolates from one patient to the same fold, so that the performance estimates are not affected by data leakage; prevalences and carrier numbers are, however, affected. Wherever a clinical recommendation is made in the main text, the deduplicated value is reported.

**Supplementary Note Table S1.** Effect of de-duplication on prevalence and performance.

|  | Ciprofloxacin | Meropenem | Ceftazidime | Tobramycin |
| --- | --- | --- | --- | --- |
| Resistance — full collection | 19.2 % | 11.9 % | 13.9 % | 8.3 % |
| Resistance — first isolate | 12.1 % | 5.8 % | 7.6 % | 4.6 % |
| Resistance — most resistant isolate | 15.7 % | 9.4 % | 12.0 % | 6.3 % |
| Brier skill score — full | 0.535 | 0.230 | 0.250 | 0.019 |
| Brier skill score — first isolate | 0.373 | 0.205 | 0.205 | 0.053 |
| Brier skill score — most resistant | 0.416 | 0.231 | 0.256 | 0.084 |
| Sens/spec at 50 blocks — full | 81.8 / 86.9 | 64.8 / 87.5 | 43.4 / 91.1 | 42.3 / 84.4 |
| Sens/spec at 50 blocks — first isolate | 86.3 / 67.3 | 94.8 / 52.5 | 46.0 / 83.2 | 77.9 / 53.3 |

### Supplementary Note 2. Reference choice and the two elongation loci Filtering against susceptible reference panels

Recoding each position against the population majority allele proved more robust than filtering against a panel of susceptible reference strains, because such a panel can exclude only those lineages that happen to be included in it. In the dataset of Cortés-Lara and colleagues [9], *mexZ* G195D passed the filter in 16 isolates, all of which belonged to ST175—a sequence type that was not represented among the 100 wild-type strains from which the filter had been created (Supplementary Figure 2c).

#### Elongation at *oprD* and *nfxB*

Protein elongation, defined as a length of more than 105% of the modal length of the locus, occurred 341 times across the 95 loci in 339 isolates; 227 of these cases concerned *oprD* and 69 *nfxB*. At OprD, elongation was associated with meropenem resistance in 49.3% of the 150 carriers, compared with 10.8% of non-carriers, corresponding to an odds ratio of 8.00. However, the same carriers also had increased odds of resistance to the other three antibiotics. Among the resistant carriers, 41% belonged to a single sequence type and 41% carried an acquired carbapenemase, whereas these characteristics were rare or absent among the susceptible carriers: 0% and 3%, respectively.

At NfxB, the univariable association was stronger: 69.2% of the 26 carriers were ciprofloxacin-resistant, compared with 19.0% of non-carriers, corresponding to an odds ratio of 9.59. Here too, however, the association was not specific: eleven of the 26 carriers belonged to one sequence type, and 16 of the 18 resistant carriers already carried a canonical *gyrA* or *parC* target mutation. When each patient and sequence type is counted only once, ten carriers remain, half of whom were resistant.

Neither of the two loci was included in the marker panel.

### Supplementary Note 3. What the specificity criterion costs

The specificity criterion penalizes mechanisms that are genuinely polyspecific. MexAB-OprM exports ciprofloxacin, ceftazidime and meropenem [30]. Therefore, any determinant that derepresses this pump should increase the odds of resistance to three of our four antibiotics. Tobramycin is not exempt, since MexXY has no outer-membrane channel of its own and functions through OprM [35,36], which is transcribed from the MexAB-OprM operon. The same argument applies to the structural genes of MexAB-OprM itself. A pump that exports three of the four agents cannot produce an antibiotic-specific marker, so its disruption enters the model as a signed feature on mechanistic grounds rather than through the specificity filter (Supplementary Note 4).

The loss of *nalD* illustrates the problem. Among the 116 isolates carrying this alteration, 62.1% were resistant to meropenem, corresponding to an odds ratio of 13.5. However, the odds were also increased for the other three antibiotics, to 12.8, 11.8 and 20.1. This results in a specificity ratio of 0.93, and the marker would be excluded. Part of the association is clearly confounded: 49 of the 116 carriers also carried an acquired resistance gene, 40 of them *bla*-IMP-1, and 41 belonged to a single sequence type. However, the effect does not disappear when the confounding is removed. After excluding the carriers of acquired enzymes, 67 isolates remained, with a meropenem resistance rate of 34.3% compared with 9.6% in the reference group. A truncating variant in the same gene, observed in 86 isolates and enriched for neither the acquired enzyme nor the sequence type, was retained with a ratio of 3.95 (95% CI 2.33–6.71). Again, the effect does not disappear when the confounding is removed.

Of the 137 marker blocks that meet the determinant criterion at one of the 33 loci with an established association with the respective antibiotic, 98 (72%) also meet the specificity criterion. For 25 of the 39 that do not meet it, the reason is not necessarily biological: the preference for one antibiotic is present, but too few isolates carry the block for the confidence bound to exclude chance; a larger collection would include these blocks.

Loss of the *mpl* gene product is the clearest example of that group. Mpl produces the pentapeptide that acts as the repressor signal for AmpR, the regulator of *ampC*. When Mpl is lost, the cell shifts towards producing more beta-lactamase. Disruption of *mpl* is an established cause of chromosomal beta-lactamase hyperproduction in clinical isolates of this species [37,38].

Ninety-seven isolates had lost the product. Of these, 46.4% were ceftazidime-resistant, compared with 13.4% of isolates that had not (odds ratio 5.61). The carriers came from 35 different sequence types and 52 patients, so the signal is not driven by a single lineage. After de-duplication, 31 carriers remained, and 41.9% of them were resistant.

The effect also holds when isolates are compared only within the same sequence type: after de-duplication, the stratified odds ratio is 11.23 across 18 strata. It rises to 14.21 once isolates with a loss in *ampD*, *ampC* or *ampR* and those carrying an acquired enzyme are removed. The signal is therefore not simply working through the loci already included in the panel (Supplementary Note 6).

The block is excluded because of insufficient specificity, not because the association is weak. Its antibiotic-specificity ratio is 1.38 (95% CI 0.85-2.24): the preference for ceftazidime is visible, but with only 97 carriers the confidence interval does not exclude chance. A larger collection would be expected to admit this block.

### Supplementary Note 4. The antibiotic-dependent direction of the pump-disruption effect

Inactivation of the MexAB-OprM and MexEF-OprN structural genes is associated with more rather than less tobramycin resistance, with stratified odds ratios of 3.97 and 4.04, respectively. The association with MexEF-OprN disappears once isolates with a disrupted mexZ or armZ (which regulate MexXY, the pump that exports aminoglycosides) are removed. This is not the case for MexAB-OprM: among 261 isolates with a disrupted mexA or mexB and intact mexZ, armZ, amgRS, fusA1 and parRS, 18.8 % were tobramycin-resistant compared with 5.3 % in isolates with none of these disruptions (stratified odds ratio 5.89). Acquired enzymes do not explain this either, since they were present in five of the 392 carriers, and neither does the loss of the shared outer-membrane channel, since oprM is intact in 328 of them. It has been shown that in a MexAB-deficient background, MexXY takes over part of the export load, and MexXY is the only one of the four pumps that exports aminoglycosides [36]. Increased MexXY production could therefore increase the tobramycin MIC without a mutation in its repressors being necessary. Two features, however, call for caution: the association is restricted to Centre A, which contributed 333 of the 392 carriers, and the MIC distribution of the carriers is bimodal rather than shifted, with an excess both at the lowest tested concentrations and at 4 to 8 mg/L.

The same reasoning extends to ciprofloxacin, because loss of MexAB-OprM also derepresses MexCD-OprJ and MexEF-OprN [39] and both export fluoroquinolones. It does not extend to ceftazidime. MexAB-OprM is the only one of the four that does, so losing it should if anything lower the ceftazidime MIC. We observe the opposite. Among carriers of a *mexA* deletion without an acquired enzyme, 26.1 % of 92 isolates were ceftazidime-resistant against 11.1 % of the remainder. This observation remains unexplained; distinguishing regulator-independent compensation from missense changes we cannot interpret would require transcript quantification.

MexXY functions with OprM, which is transcribed from the *mexAB-oprM* operon that is repressed by NalD [26,35]. Derepression should therefore increase tobramycin resistance without a mutation in the MexXY regulators being present. This is consistent with the observed association. Among isolates with intact *oprM*, no acquired aminoglycoside-modifying enzyme and no resistance-associated marker in *mexZ, armZ, amgRS*, *fusA1* or *parRS*, tobramycin resistance was 12.9 % among the 31 isolates in which *nalD* was absent (4 of 31) and 12.7 % among the 63 isolates in which *nalD* was truncated (8 of 63), compared with 2.2 % among the remaining 3,504 isolates. The two comparisons therefore give almost the same crude odds ratio, 7.1 and 6.7, with p = 0.005 and p = 0.0001, respectively. The difference in the p-value reflects the difference in group size.

Two reservations apply. It has been reported that MexXY can also work with outer-membrane channels other than OprM, several of them still unidentified [40], so the availability of OprM need not be the only route. In addition, among the isolates without *nalD*, the 61 in which *oprM* was also called absent were 91.8% tobramycin-resistant and were dominated by a single *bla*-IMP-1 clone (ST1047, 41 isolates).

### Supplementary Note 5. Where the two thresholds sit

For ciprofloxacin, meropenem and tobramycin, the group below the 5% predicted-risk threshold has an observed resistance rate below 5%. Predicted and observed resistance agree to within approximately one percentage point, and the group comprises between one-third and one-half of all isolates.

The upper threshold is a compromise. We set it at 80 % because that is the highest value at which more than one antibiotic still assigns an appreciable number of isolates to the resistant report; a higher threshold would empty that report for most drugs. The groups it produces are small, at 0.2 % to 6.8 % of isolates, so their calibration is less certain, and because the value was chosen on this collection rather than derived from an external requirement, it is the threshold most likely to move on an independent set. We therefore regard the resistant half of the report as provisional until that test has been made.

Catalogue size was examined separately. Going from the 33 loci with an established mechanistic link to all 76 lowered the cross-validated Brier skill score for three antibiotics — ciprofloxacin from 0.541 to 0.535, ceftazidime from 0.273 to 0.250 and tobramycin from 0.024 to 0.019 — and raised it for meropenem, from 0.215 to 0.230. All four figures include the signed pump feature; what that feature contributes on its own, with the catalogue held at 76 loci, is the rise from 0.220 to 0.230 given in the Results (paired difference +0.010, 95 % CI +0.005 to +0.016). In a separate analysis outside the primary model, replacing MexAB-OprM in the single signed-feature slot by a feature for MexXY disruption raised the tobramycin score from 0.019 to 0.049, by 0.030 (paired difference, 95 % CI +0.015 to +0.054); the confidence interval around the resulting score still includes zero, so tobramycin does not clearly exceed the prevalence baseline.

For ciprofloxacin, a second probability adds information that is not captured by the binary resistance category. A separate model for high-level resistance, defined as an MIC of at least 4 mg/L, was equally well calibrated and placed 77.4 % of isolates below 5 %, a group in which 1.7 % were in fact highly resistant. The report then answers two questions rather than one: whether the antibiotic will fail, and whether it will fail outright.

### Supplementary Note 6. Matched analysis within sequence types

A determinant should separate resistant from susceptible isolates within a lineage, not only between lineages. We therefore repeated the association analysis with the sequence type as a stratum, using only those sequence types that contain both resistant and susceptible isolates, and retaining one isolate per patient and sequence type. The Mantel-Haenszel estimator over these strata is equivalent to the estimate obtained from conditional logistic regression with sequence type as the stratum, but it uses every isolate within a lineage rather than arbitrarily formed pairs. Significance was assessed by the Mantel-Haenszel chi-square with continuity correction and a Bonferroni threshold over the blocks tested.

The design recovers the canonical determinants without any help from the specificity filter. For ciprofloxacin, *gyrA* position 83 reaches a stratified odds ratio of 161.8 across 56 informative sequence types (P = 6 x 10-123), *gyrA*87 reaches 32.4 and *gyrB*466 reaches 14.0. For meropenem, the surviving blocks are loss of *oprD* and *nalD*; for tobramycin, they include loss of *mexZ*. Almost everything that survives is a loss-of-function call.

For ceftazidime the surviving blocks are loss of PBP4 (145.6 across 10 strata), truncation and loss of *ampD* (17.9 and 22.5), a premature stop codon in *mpl* (13.3 across 17 strata), and loss of *nalD* (8.4). Calls at *oprD* also survive, but they reach two to four times higher stratified odds ratios for meropenem than for ceftazidime, and no transport route for ceftazidime through this porin is described.

The loss of the *mpl* gene product was examined in more detail, because Supplementary Note 3 identifies it as a marker that was set aside by the specificity criterion, not because it lacked specificity, but because there were too few carriers. For ceftazidime, it showed a clear association with resistance across all isolates. When the analysis was restricted to isolates without loss of function in *ampD, ampC,* or *ampR,* this association became even stronger. It increased further when isolates carrying acquired resistance enzymes were also excluded (P = 5 x 10^-7^, 29 carriers). This suggests that the effect is not explained by loci already included in the panel, but is instead independent of them. The same marker was also associated with meropenem resistance, although more weakly (P = 0.003). No comparable analysis was possible for ciprofloxacin or tobramycin, because *mpl* is not part of their candidate panels.

Nineteen of the 95 loci at which variants were called have no documented link to any of the four antibiotics and were therefore not assigned to a candidate panel: the peptidoglycan-precursor enzymes *murA to murG, glmM, glmS, glmU, ddlA, ddlB, mraY, alr* and *dadX,* together with *algU, mucA, mexQ* and *phzS*. They were screened nonetheless. Recoded against the population majority allele and scored for loss of the gene product in the same way as the panel loci, they yield 5,933 marker blocks. Seventy of these meet both the determinant and the antibiotic-specificity criterion for one of the four agents — 50 for ciprofloxacin, 14 for tobramycin, 5 for ceftazidime and 1 for meropenem.

Three of the seventy survive the matched analysis described above, and all three meet the antibiotic-specificity criterion for ciprofloxacin and are overlapping blocks at *mucA*: *mucA*129 (stratified odds ratio 6.59 over 10 strata), *mucA*128 (5.31 over 14) and *mucA*123 (4.23 over 15). None of them, however, explains the ciprofloxacin-resistant isolates that the panel leaves unexplained. Among the 155 ciprofloxacin-resistant isolates without a canonical target marker, 18.7 % carry a *mucA* variant, against 12.4 % of the 4,632 susceptible isolates (odds ratio 1.65, p = 0.03) — a difference far too small to account for the missing signal. For tobramycin the same variants are much more strongly enriched among the unexplained resistant isolates (45.3 % against 12.9 %, odds ratio 5.58), which is more consistent with patient-or lineage-associated sampling than with a resistance mechanism. Since *mucA* has no described role in antibiotic resistance, it was one of the 19 loci which did not contribute a marker to any result reported in the main text.

**Supplementary Table 1.**
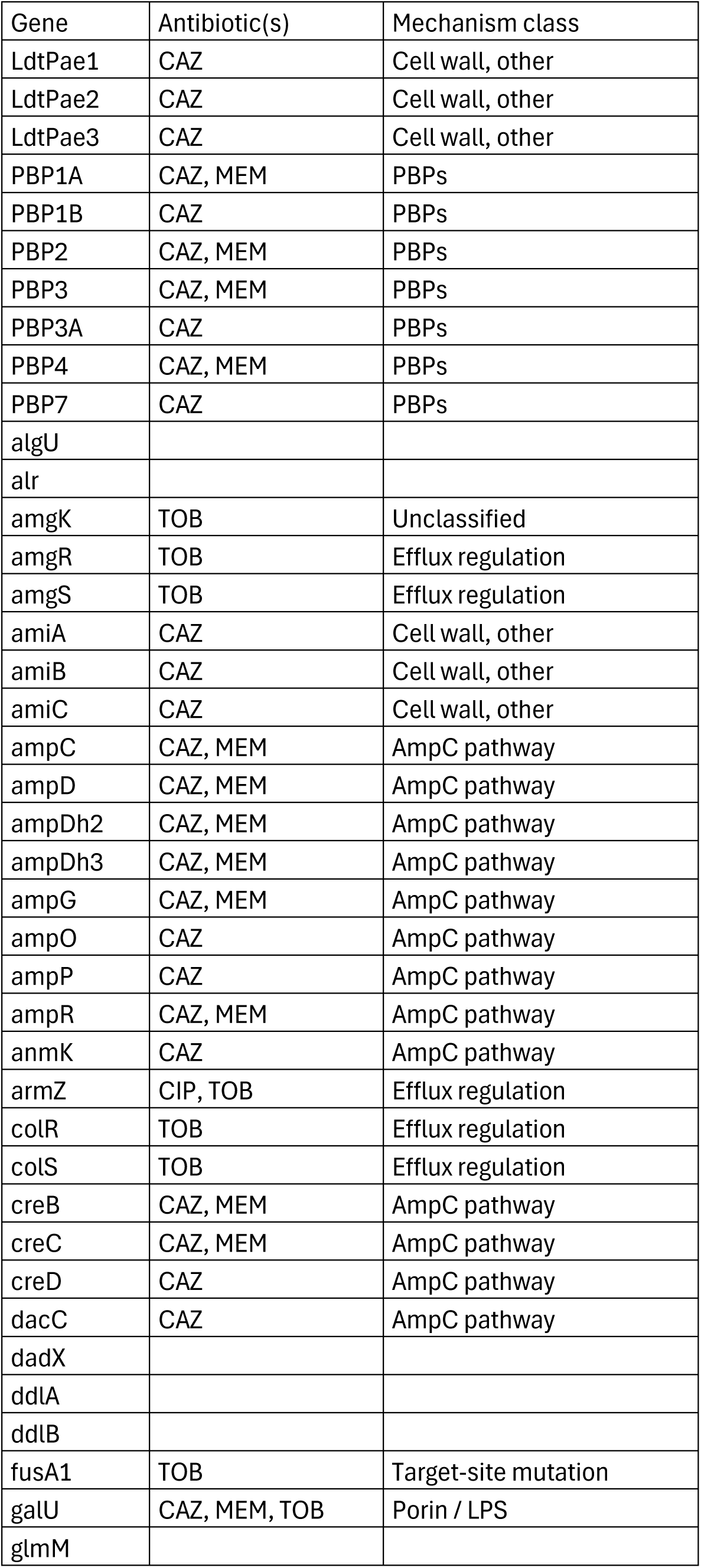
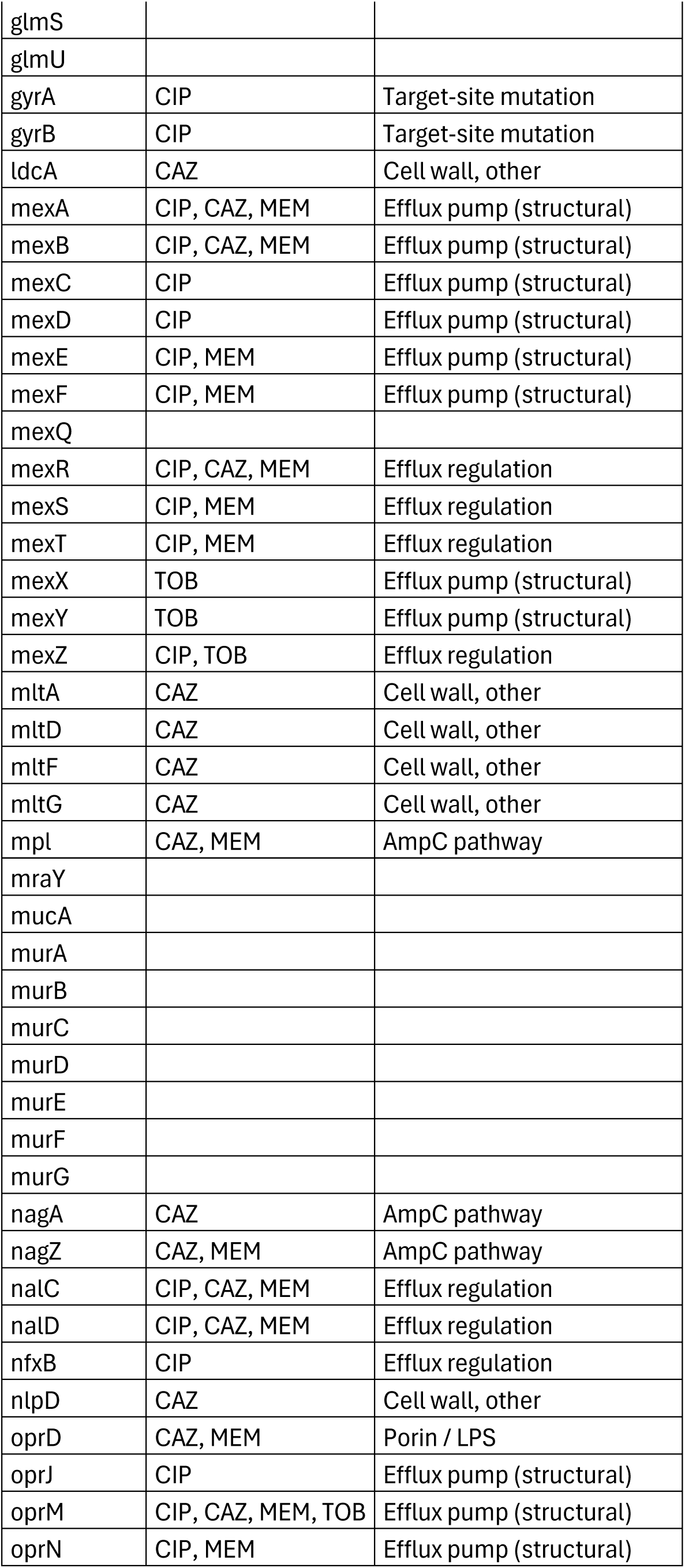
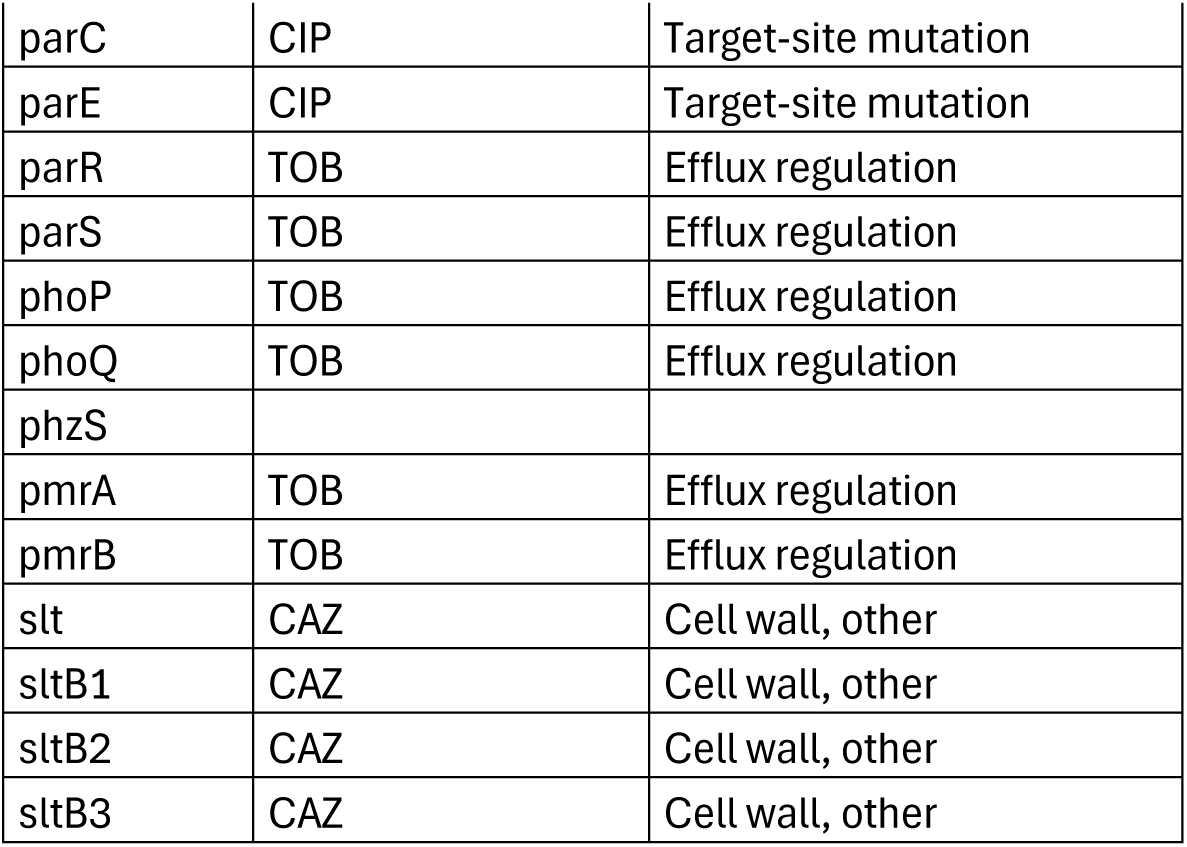

